# Vaccination strategies in structured populations under partial immunity and reinfection

**DOI:** 10.1101/2021.11.23.21266766

**Authors:** Gabriel Rodriguez-Maroto, Iker Atienza-Diez, Saúl Ares, Susanna Manrubia

## Abstract

Optimal protocols of vaccine administration to minimize the effects of infectious diseases depend on a number of variables that admit different degrees of control. Examples include the characteristics of the disease and how it impacts on different groups of individuals as a function of sex, age or socioeconomic status, its transmission mode, or the demographic structure of the affected population. Here we introduce a compartmental model of infection propagation with vaccination and reinfection and analyse the effect that variations on the rates of these two processes have on the progression of the disease and on the number of fatalities. The population is split into two groups to highlight the overall effects on disease caused by different relationships between vaccine administration and various demographic structures. We show that optimal administration protocols depend on the vaccination rate, a variable severely conditioned by vaccine supply and acceptance. As a practical example, we study COVID-19 dynamics in various countries using real demographic data. The model can be easily applied to any other disease and demographic structure through a suitable estimation of parameter values. Simulations of the general model can be carried out at [this interactive webpage](https://mybinder.org/v2/gh/IkerAtienza/SIYRD/main?urlpath=%2Fvoila%2Frender%2FSimulator.ipynb) [1].

**Author summary:** Vaccination campaigns can have varying degrees of success in minimizing the effects of an infectious disease. It is often very difficult to assess *a priori* the importance and effect of different relevant factors. To gain insight into this problem, we present a model of infection propagation with vaccination and use it to study the effects of vaccination rate and population structure. We find that when the disease affects in different ways distinct population groups, the best vaccination strategy depends non-trivially on the rate at which vaccines can be administered. The application of our analysis to COVID-19 reveals that, in countries with aged populations, the best strategy is always to vaccinate first the elderly, while for youthful populations maximizing vaccination rate regardless of other considerations may save more lives.

## Introduction

Infectious diseases have profoundly shaped human habits, and societies at large [2]. Their tremendous effects on morbidity and mortality could only be counteracted with the advent of massive vaccination campaigns, which stand out as the most effective medical interventions ever [3]. This is so despite the fact that only smallpox has been eradicated (also rinderpest in animal hosts), while multiple other communicable diseases persist at low incidence or with significantly milder effects in infected individuals – thanks to vaccination. Eradication of pathogens is difficult if they have alternative species as reservoirs, in the absence of a vaccine, or if the disease is geographically spread or has a complicated diagnosis.

Nowadays, many infectious diseases remain endemic even if a vaccine is available and its administration is widespread. Vaccinated individuals can become infected because vaccines only confer partial protection [4]; if immunity wanes, both vaccinated and previously infected individuals can get the infection again [5]. Around 1% of infected individuals undergo reinfection by SARS-CoV-2 [6], a percentage that is higher in the case of other respiratory diseases, such as influenza, where the high variability of viral strains and continued reinfections during each flu-season have made it endemic [7]. Indeed, the emergence of mutant forms of pathogens that escape immune attack and thus limit the protective effects of past infections or vaccination is the rule [8], especially when prevalence is high [9].

The eventual fate of an emergent epidemic is uncertain as it depends on the type of immunity people acquire through infection or vaccination and on how the virus evolves [10]. The effects of disease in populations along typically lengthy transients between the emergence of a contagious disease and its dynamical stabilization depend on features of the population and of the disease, but also on controlled interventions such as non-pharmaceutical measures and, critically, vaccine roll-out [11].

It is hard to quantify all possible effects of vaccination [12], since it is ideally aimed at fulfilling several dissimilar goals, among others to prevent the disease and reduce its severity and mortality and to minimize the impact of the disease on the health care system and the economy [13]. Actually, vaccination strategies often focus on one or another priority: for instance, on vaccinating first the most vulnerable groups, or on starting with the individuals with the highest spreading potential, with the hope that the most vulnerable group will be indirectly protected [14, 15]. Vaccination strategies cannot be unique and universal [16], since their effects vary as a function of the transmission mode and severity of the specific disease, of population habits (which determine contacts and therefore contagion probability), of demographic profiles and, last but not least, on the availability of vaccines [17].

In this contribution, we address the effect of different vaccination protocols in structured populations by means of a compartmental model, and assuming a non-negligible fraction of reinfections and infections of vaccinated individuals. As a measure of the effects of vaccination we use the reduction in the number of deaths after one year, as compared with a situation of no vaccination. The model is deliberately simple to focus on the overall effects of two fixed empirical variables (the infection fatality ratio, IFR, of the disease and the demographic structure of the population) and one controllable variable: the vaccination rate. In the first part of the work, we explore the effects of these variables in synthetic populations structured into two groups where the first one has a higher number of intra-group contacts (and thus is the group that best spreads the disease) and the second one contains more vulnerable individuals (with a higher IFR). In the second part of the work, we apply the model to COVID-19 and several different countries. In each particular case studied, we use available data on demography, contact matrices between age-groups [18, 19], and independently evaluated, age-dependent IFR. Finally, we have built a [public webpage](https://mybinder.org/v2/gh/IkerAtienza/SIYRD/main?urlpath=%2Fvoila%2Frender%2FSimulator.ipynb) [1] where multiple additional demographic profiles can be chosen and model parameters arbitrarily changed, also with the goal of allowing quantitative comparison with other models in the literature [14, 20].

## Models

### SIYRD: A compartmental model with reinfections and vaccination

The basic SIYRD model is a compartmental model with five different classes: Susceptible (S), Infected (I), Reinfected (Y), Recovered (R) and Dead (D) individuals. The recovered class merges individuals from two origins: those which were infected and have overcome the disease, thus bearing partial immunity to new infections, and susceptible individuals which have been vaccinated. As a first approximation, we assume that the degree of immunity acquired by vaccinated individuals is equivalent to that of recovered individuals after natural infection. Figure 1 summarizes the transitions allowed between compartments. Note that only vaccination of susceptible individuals is considered, due to the assumed equivalence between vaccination and disease overcoming.

**Fig 1.**
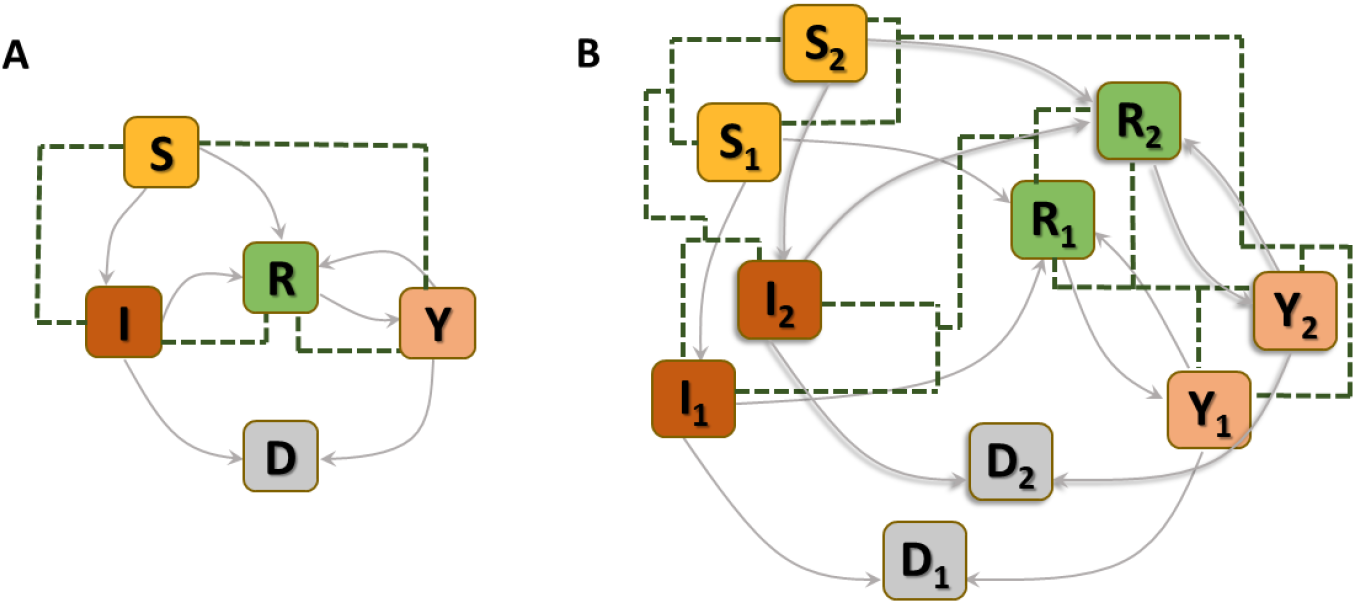
Schematic of compartments in the epidemic model with vaccination and reinfection. (A) Basic SIYRD model. Susceptible individuals (S) exit the compartment either through infection (to class I, at rate *β*_*SI*_ if infected by an I individual and at rate *β*_*SY*_ if infected by a Y individual) or vaccination (to class R, at rate *v*). Infected individuals may recover (R, rate *r*) or die (D), regardless of whether they have primary infections (I, rate *μ*_*I*_) or reinfections (Y, rate *μ*_*Y*_). Recovered individuals can become reinfected (by class Y at rate *β*_*RY*_ or by class I at rate *β*_*RI*_), and either recover again (to class R, rate *r*) or die (to class D, rate *μ*_*Y*_). Green dashed lines link classes whose interaction triggers transitions. (B) As in (A), for the model with two groups. Note that transitions occur only within classes of the same group, while interactions intra- and inter-groups (green dashed lines) couple the dynamics. These interactions are weighted through contact matrices and the fraction of individuals in each group, which consider the demographic structure of the population.

The equations that define the epidemic model are

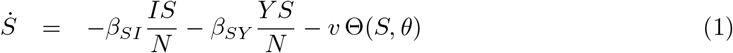

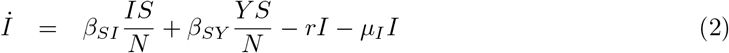

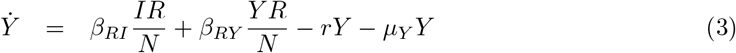

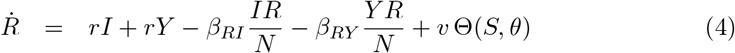

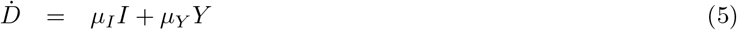

Parameters are rescaled in such a way that the time unit of our simulations is one day. Vaccination is implemented in this model through a parameter *v* that represents the fraction of population vaccinated per time unit, i.e. per day. This rate is multiplied by a function Θ(*S, θ*) that takes into account its progressive slowdown and eventual halt when a fraction *θ* of individuals has been vaccinated. The specific functional form of Θ(*S, θ*) does not affect the overall dynamical properties of the system (see S3 Appendix).

#### Model parameters

The separation between primary infections (I) and reinfections (Y) entails four different infection rates (see Fig. 1): *β*_*SI*_ and *β*_*SY*_ correspond to the infection rates of susceptible individuals by primary-infected and re-infected individuals, respectively; *β*_*RI*_ and *β*_*RY*_ are the equivalent infection rates for recovered individuals. Although the precise values of some of these infection rates might be difficult to estimate in general, we consider disease types where they are subject to certain relations that hold on average, over the population. For example, reinfected individuals have a lower infective capacity in comparison to primary-infected individuals, both towards susceptible and recovered individuals. In other words, reinfection rates are smaller than primary-infection rates, *β*_*SI*_ ≥ *β*_*SY*_ and *β*_*RI*_ ≥ *β*_*RY*_.

Consistently, the likelihood that a susceptible individual becomes infected is larger than that of a recovered individual, since the latter bears at least partial immunity against the disease either due to a prior infection or to vaccination. This applies both to primary infections, *β*_*SI*_ ≥ *β*_*RI*_, and to reinfections, *β*_*SY*_ ≥ *β*_*RY*_. Finally, we assume that the ratio between the infection rates of primary-infected and reinfected individuals is independent of the state of the individual that can be potentially infected,

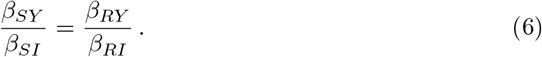

The mortality rate of primary-infected individuals can be calculated using the infection fatality risk (IFR) of the disease under consideration, defined as the ratio between the number of fatalities and the number of infections in a given (sub)population (see S2 Appendix). The mortality rate of reinfected individuals is assumed to be significantly lower, *μ*_*I*_ *≫ μ*_*Y*_, as a result of their partial immunity against the disease. Finally, for simplicity we assume that the recovery rate *r* of primary-infected and reinfected individuals is identical, neglecting possible small differences.

The quantitative estimation of *r* depends on the IFR and on the infectious period of the disease, see S2 Appendix. The vaccination rate *v* is a variable that can be explored in a range of values depending on vaccine supply. We will consider values up to 2%, representing the fraction of population vaccinated in one day. A summary of parameters in the SIYRD can be found in Table 1.

**Table 1.** Definition of parameters in the SIYRD model. All rates have dimensions of day^−1^ with the exception of the vaccination rate, which has dimensions of population (fraction) *×* day^−1^, and *θ*, which has dimensions of population (fraction).

| Parameter | Definition |
| --- | --- |
| $\beta_{SI}, \beta_{RI}$ | Infection rate of S and R individuals by I individuals |
| $\beta_{SY}, \beta_{RY}$ | Infection rate of S and R individuals by Y individuals |
| $r$ | Recovery rate of I and Y individuals |
| $\mu_I$ | Mortality rate of I individuals |
| $\mu_Y$ | Mortality rate of Y individuals |
| $v$ | Vaccination rate |
| $\theta$ | Maximum fraction of vaccinated population |

### Stratified SIYRD (S^2^IYRD)

In order to consider the demographic structure of populations and how a given disease affects different groups (e.g. depending on age or sex), we extend the SIYRD model to represent two different population groups (*G*_1_ and *G*_2_) per class, each containing *N*_1_ and *N*_2_ individuals, respectively. Variables and parameters characteristic of each group now get the corresponding subindex: this applies to vaccination rates *v*_*i*_, mortality rates 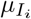 and 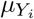, and recovery rates *r*_*i*_. Disease transmission rates *β*_*SI*_, *β*_*SY*_, *β*_*RI*_ and *β*_*RY*_ are independent of the group to which they apply. To be consistent, the two groups are connected to each other through a contact matrix that reflects individual connectivity patterns (see S1 Appendix). To simplify notation, the simple, one-class model with reinfection and vaccination will be called SIYRD all through the text, while the model with two coupled groups will be called S^2^IYRD.

Equations (7-11) describe the dynamics of group *G*_1_ in the S^2^IYRD model,

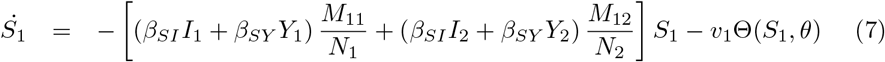

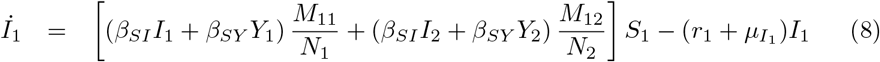

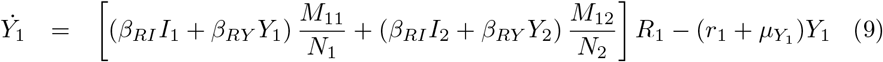

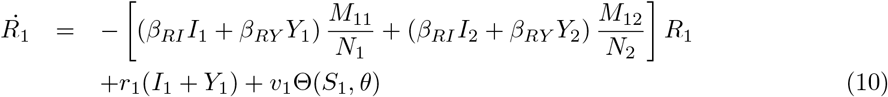

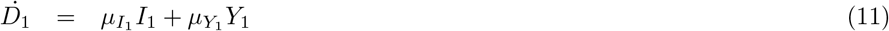

This model has a second set of analogous equations, coupled to Eqs. (7-11), obtained by simply interchanging subindexes, 1 ↔ 2.

The most important modification of the model comes from the matrix of contacts **M**, whose elements *M*_*ij*_ represent the number of contacts an individual of group *i* has with individuals of group *j*, thus weighing the effect of intra- and inter-group contacts in contagion rates (see S1 Appendix).

#### S^2^IYRD model parameters

Though mortality and recovery rates are in principle subject to relationships analogous to those described for the SIYRD model, they can be also independently estimated for each group as characteristics of the disease under consideration. Now, the values of the elements *M*_*ij*_ synthesize the contact structure of the affected population and, in general, can be empirically obtained using actual demographic structure and surveys of contacts between different groups in a variety of daily situations (see S1 Appendix). Finally, the vaccination rate can be varied to represent different administration protocols, in order to test how they affect disease progression. We will consider three situations in which priority is given to *G*_1_, to *G*_2_, or both groups are simultaneously vaccinated. The result of each protocol will be compared with the baseline of no vaccination. Specifically, the prioritized population group is vaccinated at a rate *v* until the fraction of susceptible individuals reaches (1 − *θ*). When that happens, vaccination of the second, non-prioritized group begins, provided the previous threshold *θ* has not been yet reached through natural infections. Under simultaneous vaccination, both groups are vaccinated at rates proportional to their respective group sizes. If vaccination finishes in one of the groups first, the other group receives all doses. Note that, due to the vaccination prescription, where a fixed number of individuals (equal to the vaccine doses available) are vaccinated at each time, our simulations will be performed with finite populations of size *N* = 10^8^, and with the real population size in specific examples. To permit comparison of different results, however, we will use normalized variables *n*_*i*_ = *N*_*i*_*/N*.

In every specific scenario, three factors emerge as the main determinants of epidemic spread and thus of optimal vaccination protocols: the division of the population into groups and their contact habits (subsumed under the *M*_*ij*_ terms), the aetiology of the disease (IFR and recovery, death and infection rates), and the vaccination rates *v*_*i*_ —which stand as the only variables that can be externally modified when the disease propagates freely or under constant non-pharmaceutical measures.

## Results

### Dynamics of the SIYRD model

#### Steady states

The linear stability analysis of the SIYRD reveals the existence of a unique nonzero steady state corresponding to epidemic mitigation: susceptible and infectious individuals vanish, leaving only recovered and dead individuals (0, 0, 0, *R*^*^, *D*^*^). The exact values of *R*^*^ and *D*^*^ can be calculated under certain conditions (see S3 Appendix). This fixed point is always neutrally stable. If no infection of recovered individuals by reinfected ones is allowed (*β*_*RY*_ = 0), the qualitative result remains unchanged.

When vaccination is suppressed, a second steady state is also possible, (*S*^*^, 0, 0, *R*^*^, *D*^*^), which is linearly unstable and therefore irrelevant from an epidemiological viewpoint. A neutrally stable pure endemic steady state (0, 0, *Y* ^*^, *R*^*^, *D*^*^) emerges if the mortality rate of reinfected individuals is set to zero, *μ*_*Y*_ = 0. Further details on fixed points and their stability, together with figures illustrating the dynamics towards different final states can be found in S3 Appendix. In principle, it is not possible to determine to which steady state the system will converge given the initial conditions and specific parameter values.

#### Long transients with a quasi-endemic disease

Due to the introduction of reinfection, the model presents an interesting behavior in a broad range of parameters: the fraction of reinfected individuals *Y* and the total number of fatalities *D* experience a long, plateau-like transient before relaxing to their asymptotic values, see Figure 2. This behavior is related to the existence of a stable endemic state for *μ*_*Y*_ = 0. Indeed, the mechanistic origin of long transients relies on the loop that links R and Y classes, whose dynamical equations fully decouple from the rest in the limit when *S* = *I* = 0 and *μ*_*Y*_ = 0, and present slow dynamics for small values of *μ*_*Y*_. In other words, these long transients are mostly controlled by the low mortality rate of secondary infections, which limits the flux of individuals out of the Y-R loop and delays the end of the epidemic. While, outside of this regime, the asymptotic state is reached in about a year time, it may take over a century for the disease to disappear in the slow regime. This behavior is observed for a broad range of parameters in the relevant case when the dynamics of first infections occurs at rates several-fold faster than that of secondary infections. In practice, what the model yields in a few years time is a long plateau in epidemic incidence that can be interpreted as a quasi-endemic disease. At longer timescales, however, the model is no longer a valid description of the dynamics, since variables such as immune decay, mutations yielding new viral variants and, eventually, demographic changes, come into play. It is in these situations when diseases can become truly endemic, since the pool of susceptible individuals is replenished through one or another mechanism.

**Fig 2.**
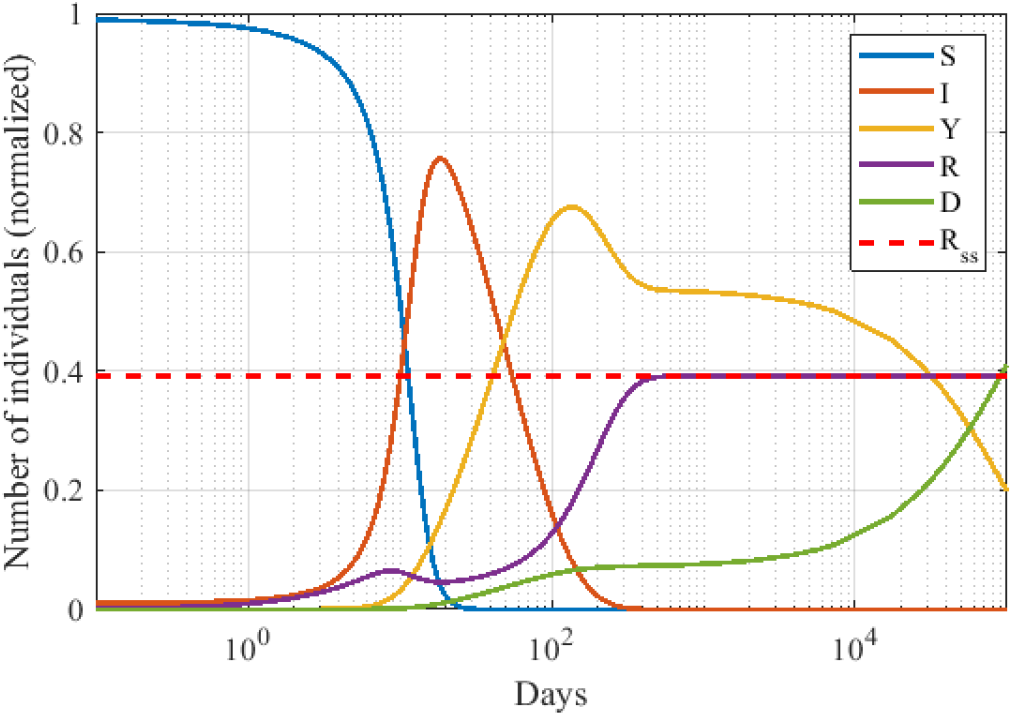
Generic dynamics of the SIYRD model. The existence of reinfection causes a non-monotonous increase in the number of recovered individuals. After an initial increase due to the I → R transition, this fraction decreases when I grows further since reinfections thus become more frequent. For small values of the death rate of Y individuals, *μ*_*Y*_, a long plateau in the number of secondary infections develops, significantly delaying the extinction of the epidemic (note the logarithmic *x*−axis). Parameters are *β*_*SI*_ = *β*_*RI*_ = 0.46, *β*_*SY*_ = *β*_*RY*_ = 0.046, *μ*_*I*_ = 0.00149, *μ*_*Y*_ = 10^−5^, *r* = 0.0180, *v* = 0.01, and *θ* = 0.7. The red dashed line represents the asymptotic value of *R*^*^ in a case where it can be analytically calculated (see S3 Appendix for further details). Initial conditions: (0.99, 0.01, 0, 0, 0).

### Dynamics of the S^2^IYRD model

In this section, we will discuss the dynamics of the S^2^IYRD model under the three possible vaccination strategies: priority to *G*_1_, to *G*_2_, or simultaneous vaccination. The advantage of either strategy will be evaluated under variable vaccination rates. Here, we do not make explicit the criterion to split the population into two groups, and explore instead the effect of these two groups having dissimilar sizes and being differently affected by the disease. To this end, we will study two representative values of the contact strength between groups, vary the fraction of population in either group and the IFR ratio, and illustrate how these affect the optimal vaccination strategy under increasing vaccination rates. Parameters can in principle be varied independently, but the closure relationship *M*_12_*n*_1_ = *M*_21_*n*_2_ (see S1 Appendix) has to be fulfilled for consistency. Also, we will restrict our explorations to cases where the group at higher risk is the slower spreader; otherwise, there is no conflict between vaccinating the most susceptible or the group that best spreads the disease, and the best strategy is therefore trivial. Without loss of generality, group *G*_1_ will be the faster spreader, and group *G*_2_ the one most affected by the disease. We will use as measure of the effectiveness of a strategy the asymptotic decrease in the number of fatalities once the epidemic halts (or, alternatively, at a sufficiently long time since vaccination started).

#### General dynamical features

The behavior of the model is qualitatively robust for all scenarios explored, for different parameter values and contact matrices, as long as the initial reproductive number is slightly above the epidemic threshold (*R*_0_ = *β*_*SI*_*d*_*I*_, ≳ 1). Nevertheless, important quantitative differences arise (see examples in the next section). As time elapses, the number of susceptible individuals in both groups is reduced due to infection and vaccination. Simultaneously, there is an increase in the number of infected individuals and a subsequent increment of recovered and dead individuals. Furthermore, increases in the number of recovered individuals may cause a boost of reinfections under sufficiently high prevalence. Ultimately, when susceptible individuals become exhausted, primary infections decline and model dynamics become exclusively dependent on recovered and reinfected individuals. Infection rates and contact between groups mostly determine the speed of the epidemic, especially affecting how fast susceptible individuals move to other compartments. In turn, mortality and recovery rates play the largest effect in setting the final number of deaths. If the previous parameters are fixed, it is the vaccination rate that determines the temporal evolution of the epidemic and the optimal vaccination strategy.

#### Effect of model parameters in total fatalities

Let us explore several synthetic examples to illustrate important relationships between model parameters and their effect on death reduction. Figure 3 summarizes the overall behavior and illustrates how variations in the vaccination rate and in the relative size of the groups affect optimal vaccination protocols.

**Fig 3.**
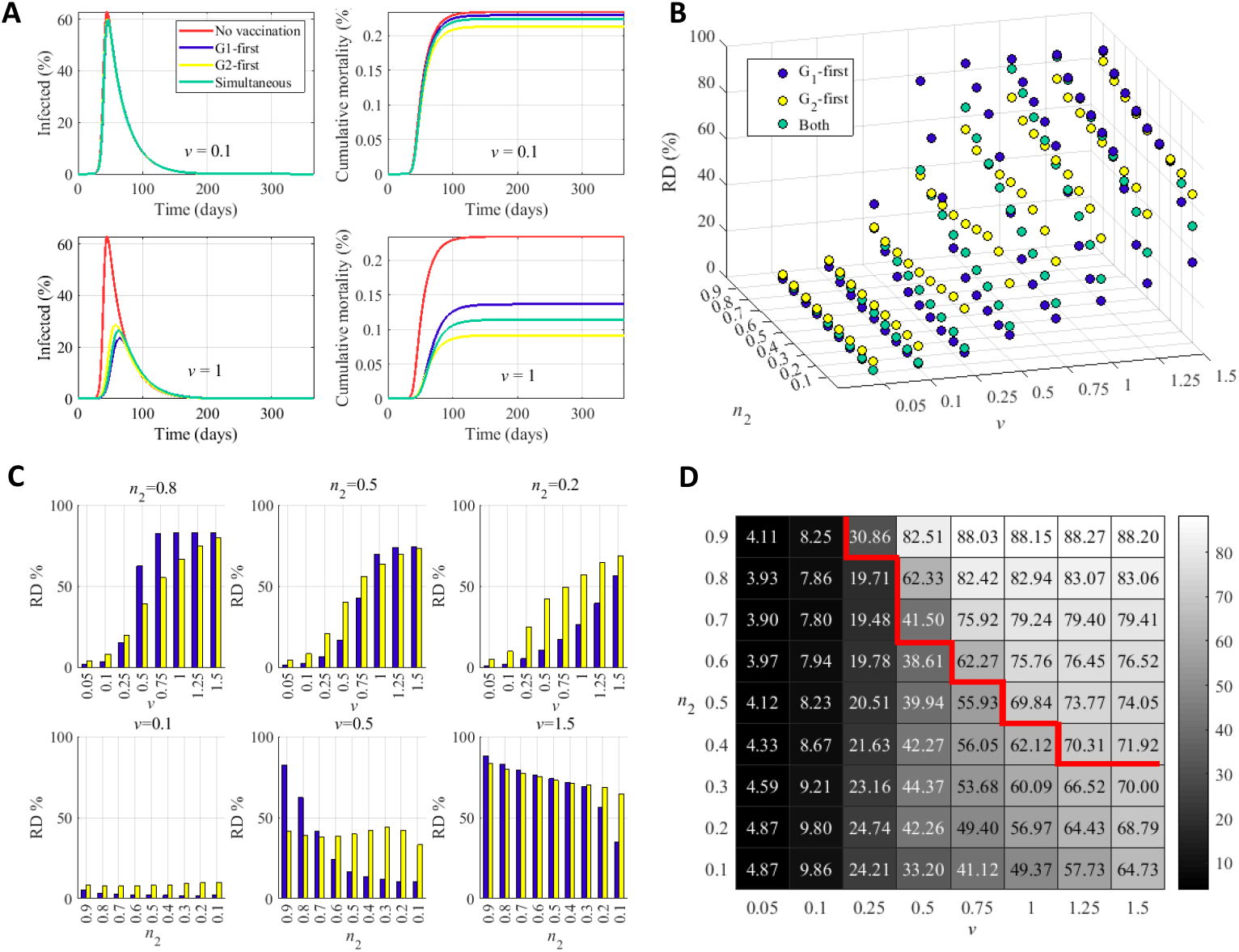
Summary of the dynamics of the model and final states as a function of vaccination rate and relative group size. (A) Dynamics of the system for low (above, *v* = 0.1) and high (below, *v* = 1) vaccination rates, with *n*_2_ = 1*/*3. (B) Reduction in the number of deaths (RD) for each strategy (see legend) as a function of the relative group size and the vaccination rate. In the absence of vaccination, RD = 0. (C) Reduction in the number of deaths for the optimal strategy. Panels stand for three different relative group sizes, as a function of the vaccination rate (above) and for three vaccination rates, as a function of the relative group size (below). Blue bars correspond to priority given to *G*_1_ as the best strategy; yellow bars correspond to priority given to *G*_2_. (D) Quantitative effect of the best strategy as a function of the vaccination rate and the relative size of the groups. To the left of the thick red line, vaccination of the *G*_2_ group first yields higher RD (numerically obtained values shown in each cell), and *vice versa* to the right of the line. Panels B, C, and D convey similar information, but underscore different effects. Parameter values common for all simulations are: *N* = 10^8^, *M*_11_ = 6, *M*_22_ = 3, *M*_12_ = 2, IFR_1_ = 0.1, IFR_2_ = 0.5, *d*_*I*_ = 15*d, r*_1_ = 0.0667, *r*_2_ = 0.0663, 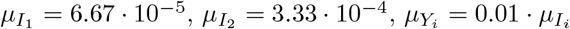, *β*_*SI*_ = 1*/d*_*I*_ = 0.0667, *β*_*RI*_ = 0.1 *· β*_*SI*_, *β*_*RY*_ = 0.5 *· β*_*SI*_, *β*_*SY*_ = *β*_*RY*_ *· β*_*SI*_*/β*_*RI*_ and *θ* = 0.7.

Characteristic dynamics under slow and fast vaccination rates are represented in Fig. 3A. If the vaccination rate is too slow, the dynamics are barely distinguishable from free propagation, and the overall reduction of the number of fatalities is very modest. For sufficiently high vaccination rates (around 0.5 and higher), the reduction of deaths becomes significant. Figure 3B represents the reduction obtained in number of deaths for each strategy as a function of the vaccination rate and the relative group size. At low vaccination rates, the advisable strategy is to vaccinate first group *G*_2_, the most vulnerable, though the optimal strategy changes to priority given to group *G*_1_ for sufficiently high vaccination rates, the precise value of *v* at which the best strategy changes depending on the relative size of the groups (see also Fig. 3C). The selection of the optimal strategy is more critical at intermediate values of the vaccination rate and when the two groups have comparable sizes. At very high vaccination rates, however, vaccination strategies yield similar reductions and are weakly dependent on the relative group size. Figure 3D presents a quantitative summary of the effect of the best strategy. Cells to the left of the thick dark red line correspond to priority to *G*_2_ as the advisable protocol, with a reduction in the number of deaths given for each vaccination rate and population fraction *n*_2_. To the right of the thick dark red line, the optimal strategy is priority vaccination of group *G*_1_.

We have explored the effect of the IFR in the optimal vaccination strategy as a function of the vaccination rate for two different group sizes (or contact matrices, recall the coupling between these two quantities through the closure relation), see Figure 4. This case serves to compare the effects of diseases of different severity (as represented by the variation in the IFR) that, however, share their propagating ability and affect the same population.

**Fig 4.**
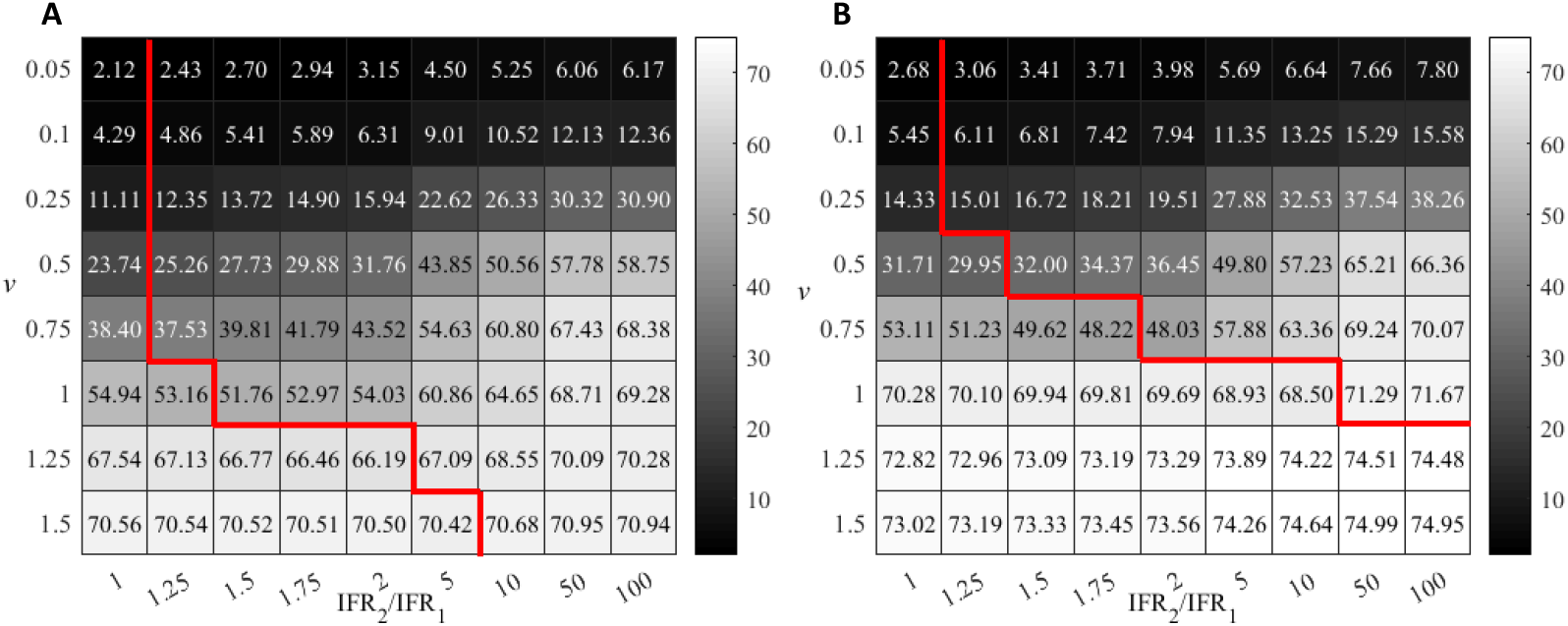
Reduction of death values (%) achieved by the optimal vaccination strategy as a function of vaccination rate and IFR ratio. Two scenarios, with *M*_12_ = 2 (A) or *M*_12_ = 1 (B) are simulated. To the left of the thick red line *G*_1_ prioritization is the optimal strategy. To the right, vaccination of the *G*_2_ group first yields the highest RD. Except for the IFR ratio, parameters are as in Fig. 3 with *n*_2_ = 1*/*3.

At low IFR ratio, the strategy yielding the higher death reduction is vaccination of the group with the higher number of contacts, *G*_1_ in our examples, since both groups experience similar effects of the disease. The situation is reversed as the IFR ratio increases, since as the disease impinges more severely on the *G*_2_ group, its priority vaccination becomes more advantageous. As in the examples above, priority vaccination of the most vulnerable group is advised for low vaccination rates. Consistently, the optimal protocol changes at sufficiently high *v*, the threshold depending on the IFR and on the difference in contacts (c.f. group sizes) between the two groups.

As the results in this section illustrate, changes in different parameters are non-linearly related, and it does note seem possible to predict the optimal strategy without taking into account all variables and quantifying their effect through explicit simulations of the epidemic model. On the one hand, as vaccination rate increases, the proportion of vaccinated *G*_2_ individuals grows even if this is not the priority group, since *G*_1_ vaccination can be anyway completed earlier, and vaccination of *G*_2_ individuals begins right after. Moreover, indirect protection of most vulnerable individuals through mitigation of *G*_1_-spreading capacity increases too. This is also reflected in the effect of reducing inter-group contacts. When *M*_12_ is reduced, *G*_1_ or simultaneous priority are more likely to improve *G*_2_ death reduction at a given vaccination rate. Eventually, the *G*_1_-priority approach often becomes the optimal strategy for sufficiently high vaccination rates.

### Application of S^2^IYRD to COVID-19

It has been proven that age is strongly correlated with severity and mortality of COVID-19 [21–27], a disease where reinfections are not rare [28–30]. Therefore, an assessment of different vaccination scenarios for SARS-CoV-2 propagation using the age-stratified S^2^IYRD model appears as a useful framework to detect major qualitative differences between vaccination strategies. In this section, we take different demographic profiles and empirically measured COVID-19 IFRs as case-examples. Populations will be divided into two groups through an age threshold. Contact matrices calculated from available data allow to estimate parameters *M*_*ij*_ for a given population split. Since there are multiple parameters that can be varied, we will discuss illustrative examples in this section and leave to the interested reader to explore further cases through the [interactive webpage](https://mybinder.org/v2/gh/IkerAtienza/SIYRD/main?urlpath=%2Fvoila%2Frender%2FSimulator.ipynb) [1] that we have developed.

Figure 5A reports empirical data on IFR per group age for COVID-19 in Spain. COVID-19 fatality risk grows close to exponentially with age and with a similar slope (multiplying by about 4 the risk per decade) in all countries analysed [31]. IFR profiles as in Fig. 5A allow to calculate the values of IFR in groups *G*_1_ and *G*_2_, and the mortality and recovery rates of primary infections (see S2 Appendix). The demographic composition of the population, as in the example shown in Fig. 5B, is needed to estimate the values *M*_*ij*_. All estimated parameters vary if the threshold age that separates both groups, which affects their composition and the intra- and inter-group contacts, changes (see Fig. 5C). In the case of Spain, relative differences between the two groups in IFR and number of contacts are maximized for an age threshold at 80 years (down-right panel in Fig. 5C). We will keep this threshold fixed in this section to focus on the relationship between demographic profiles and vaccination strategies. A second case-example, with a threshold at 50 years, can be found in S4 Appendix (again, different thresholds and various other countries, India, Italy and Japan at the time of this writing, can be explored in the [the webpage](https://mybinder.org/v2/gh/IkerAtienza/SIYRD/main?urlpath=%2Fvoila%2Frender%2FSimulator.ipynb) [1]).

**Fig 5.**
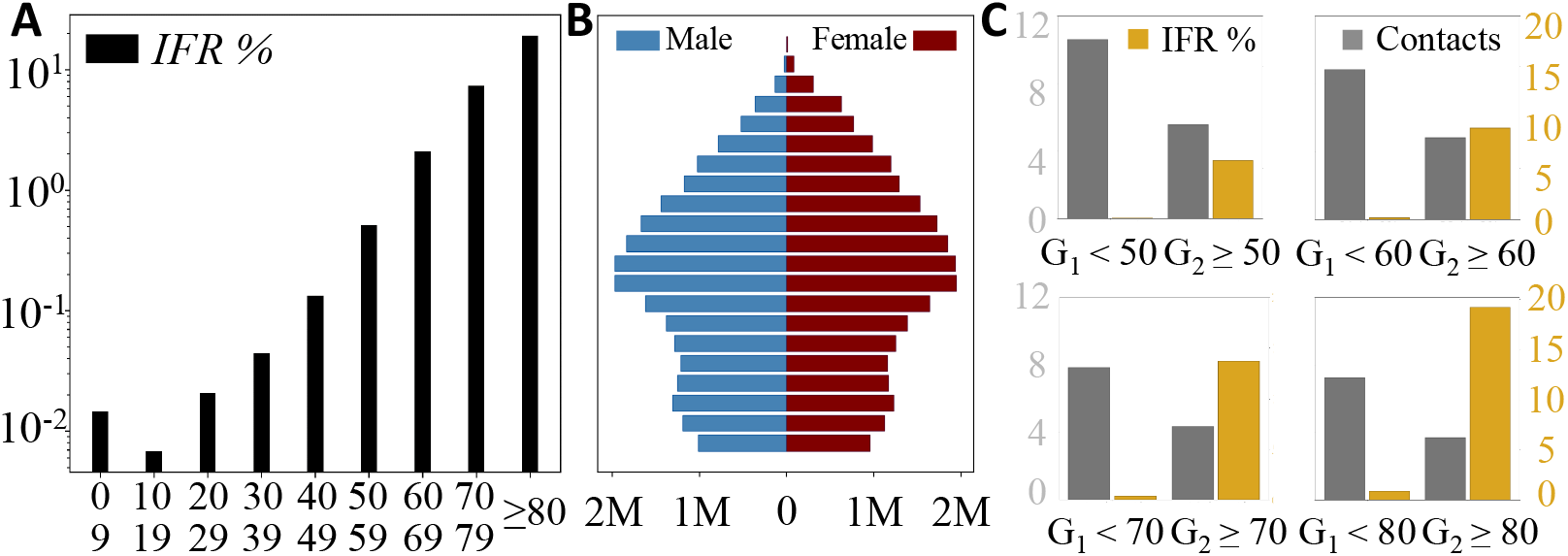
S^2^IYRD parameters for COVID-19 can be estimated from the empirical fatality risk (IFR) and demographic profiles. (A) IFR percentage values based on COVID-19 confirmed fatalities as a function of age in Spain (see S2 Appendix). (B) Demographic pyramid for the Spanish population in 2020; blue corresponds to male population, dark red to female population; in the vertical axis, populations are grouped in 5-year intervals, starting with 0-4 years at the bottom. Different age thresholds yield different values of the fraction of elder group *n*_2_, (*n*_1_ = 1 − *n*_2_). (C) In gray, effective values of the number of contacts in groups *G*_1_ and *G*_2_ and, in gold, their corresponding IFR percentage values for four different age thresholds separating both groups, as shown in the legend. See S1 Appendix and S2 Appendix for details.

Table 2 summarizes the empirical values obtained for four different countries which differ in their demographic composition and in their contacts (see S2 Appendix for raw data and other details). Spain has a negative growth in recent decades that yields a narrowing, cup-like demographic pyramid. It also bears the largest fraction of elder population, over six-fold that of the South African Republic (SAR). Israel’s demographic pyramid has the Christmas-tree-like shape characteristic of young and rapidly growing populations, with a fraction of population above 80 years between that of Spain and the SAR. Such is the case of the elder group in the USA as well. The USA demographic pyramid displays a box-like shape that reveals a population of stable size. The IFR of all four countries is comparable for the elder group but differs for the young one. In turn, the fraction of contacts within group *G*_1_ is high and comparable, while values of contacts between groups and within the *G*_2_ group vary.

**Table 2.** Empirical parameters for the countries in Fig. 6. Updated demographic pyramids (to year 2020) and independent, empirical values of COVID-19 IFR for each country per age group have been used for these estimations. Details on data origin are reported in S1 Appendix and S2 Appendix.

| | $n_2$ | IFR$_{n_1}$ | IFR$_{n_2}$ | $M_{11}$ | $M_{22}$ | $M_{12}$ |
| --- | --- | --- | --- | --- | --- | --- |
| Spain | 0.06 | 0.87% | 19.26% | 6.83 | 0.34 | 0.21 |
| SAR | < 0.01 | 2.88% | 19.85% | 6.16 | 0.26 | 0.07 |
| Israel | 0.03 | 0.38% | 18.51% | 6.79 | 0.84 | 0.09 |
| USA | 0.04 | 1.04% | 21.53% | 6.34 | 0.62 | 0.15 |

As previously done, we now examine three different vaccination strategies: priority to the over-80-years group *G*_2_, priority to the younger group *G*_1_, and simultaneous vaccination proportional to the population of either group. In the first two scenarios, vaccination of the corresponding second group starts once that of the priority group finishes (with a threshold *θ* = 0.7). Fig. 6 summarizes the effect of different vaccinating protocols in the reduction of deaths as a function of the vaccination rate *v* for the four countries above. In all cases, vaccinating first *G*_2_ always reduces mortality more than does any of the other two strategies, often more than doubling the effect of alternative protocols at low vaccination rates (see Fig. 6). Nevertheless, the difference is minor for the case of the SAR due to two main factors: the relatively high IFR of the younger group and, especially, the very low size of the elder group. As a result, most deaths come from *G*_1_, blurring the positive effect of protecting first the elder group. On the other hand, the vaccination of this latter group occurs early in the monitored year and, as explained, then continues with group *G*_1_. Therefore, its effects are not that different from simultaneous vaccination (and, actually, from priority to the *G*_1_ group) in one-year time. Still, differences are observable at the initial stages of vaccination. Remarkably, simultaneous vaccination or priority vaccination to the *G*_1_ group do not present major differences in any case. In the case of Israel, vaccination of the *G*_1_ group first is slightly beneficial at low vaccination rates, while simultaneous vaccination is more efficient at high *v*; in the case of the USA, simultaneous vaccination is better at intermediate vaccination rates.

**Fig 6.**
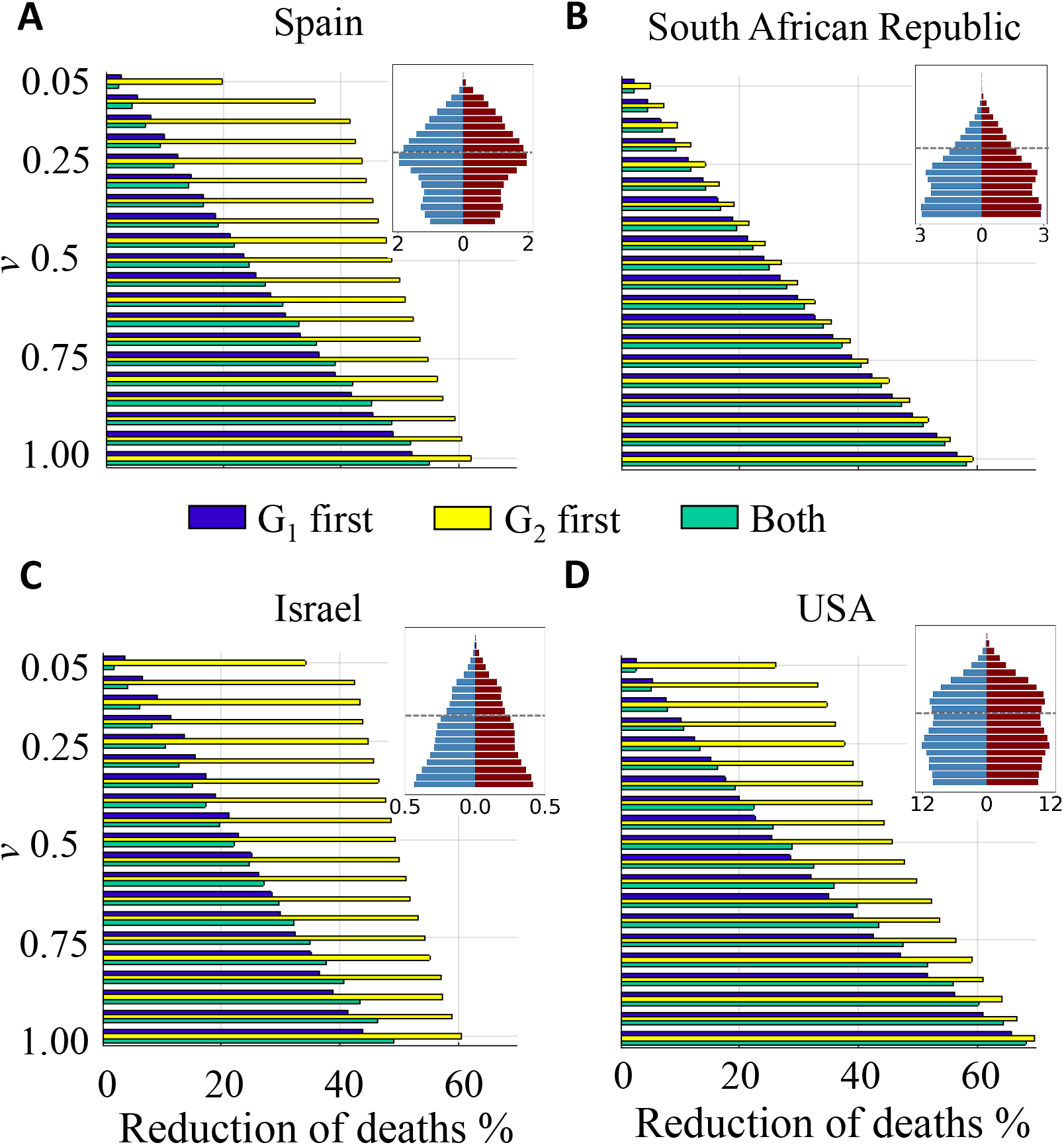
Death reduction after one year corresponding to each vaccinating strategy as a function of the vaccination rate. (a) Spain; (b) South African Republic; (c) Israel and (d) USA. Population pyramids are shown as an illustration of the demographic structure; blue corresponds to male population, dark red to female population; numbers in the x-axes of the insets stand for population in millions; in the vertical axis, populations are grouped in 5-year intervals, starting with 0-4 years at the bottom. Demographic data correspond to year 2020 and have been obtained from the Spanish National Institute for Statistics (INE) and from the World population 2019 prospects of UN. IFR measures independently carried out in each of these countries have been used in the simulations (see S2 Appendix); otherwise, model parameters are the same for all cases, with an age threshold at 80 years (indicated as a dashed line in the inset).

Vaccination rates, however, do make a significant quantitative difference in the difference of performance between the optimal strategy and the rest. In terms of reduction of fatalities, the advantage of *G*_1_ priority barely reaches a 5% with respect to the worst-performing protocol. This even requires relatively high vaccination rates (around *v* = 1), and the improvement typically decreases with decreasing *v*.

Increasing the vaccination rate monotonically increases the reduction in the number of deaths, regardless the protocol. The increase is almost linear for Spain and Israel, but accelerates with *v* for the SAR and the USA, especially. For example, if priority is given to the group *G*_1_, an increase in 0.05% in the vaccination rate from *v* = 0.2 to 0.25 rises RD in 2.3% points, while an increase from *v* = 0.9 to *v* = 0.95 causes a 4.9% increase in RD. The change in RD is also about two-fold for the other two strategies.

At sufficiently high vaccination rates, the three vaccination strategies reduce the differences in their effect. However, even in that situation, *G*_2_-prioritization allows to attain much earlier in time a death reduction comparable to that eventually attained with any of the other two strategies, due to the early protection of the most susceptible group.

## Discussion

The goal of this work has been to analyse major effects of demographic population structure, disease fatality and vaccination rate in optimal vaccination strategies in a model with reinfections, and to provide a general scenario that can be applied to other situations. An important variable whose effect cannot be disentangled from other demographic factors is the contact between groups, which affects optimal vaccination strategies through its implicit relevance in disease spread. The dynamics have been evaluated using effective population-level contact matrices estimated in situations without mobility restrictions [18]. Contact matrices are however a key element that becomes modified in presence of an on-going pandemic due to non-pharmaceutical interventions, as indirectly shown by changes in mobility [32] that directly impact propagation dynamics [33]. The model incorporates other simplifications such as the implementation of a limited number of compartments and the use of only two groups.

Optimal vaccination roll-out depends on multiple variables. The precise value of the vaccination rate at which the advantage of vaccinating first the most susceptible group is lost in front of alternative strategies sensitively varies with the IFR of the specific disease. At high IFR, as for COVID-19, priority vaccination to the most connected group is only advantageous at remarkably high vaccination rates (above 1%, the precise threshold depending on the characteristics of the population). However, the relative advantage of the optimal strategy is reduced as the number of administered daily doses grows. The reduction is sensitive to demographic differences in a non-trivial way: while in the South African Republic and in the USA our model yields a quasi-equivalence among the three different strategies for a vaccination rate *v* ≥ 1%, a significant difference persists for Spain and Israel, for example. Our results with two age thresholds (at 80 and 50 years) are consistent with the overall picture. Lowering the age threshold implies a reduction of IFR in the older group and a simultaneous increase in the average number of contacts per individual in both groups. In terms of death reduction, priority vaccination of the older group is systematically more efficient for high IFRs (as those of COVID-19). For diseases with lower IFR, as influenza, the general consensus is that children should be vaccinated first due to their high transmission capacity [17, 34]. It is likely that the optimal strategy for COVID-19 changes and resembles that for influenza if the disease becomes endemic and its effects turn milder once most of the world population has been either infected or vaccinated.

These results are consistent with those of other model-based studies showing that the optimal vaccination strategy balancing direct and indirect protection against COVID-19 is highly determined by vaccine supply and efficacy [14, 35, 36] and vaccination of the most susceptible group is more efficient under a broad range of situations [37, 38]. Our study adds two elements to previous approaches. First, the introduction of reinfections, which are non-negligible for COVID-19 and other coronaviruses [5], shows the existence of long transients (or quasi-endemic states) that may predate the transition to a truly endemic state predicted for COVID-19 [11]. Second, the model is simple enough so as to allow the characterization of systematic effects due to, at least, group size, demographic composition and IFRs.

As other models used to inform optimal vaccination roll-out [14, 35–37], our model assumes that individuals within a given group have on average the same number of contacts. More realistic models should consider heterogeneity in the number of contacts, since it has been shown that highly skewed contact distributions, with hubs (or, equivalent from a dynamic viewpoint, super-spreader individuals or events) have important effects in immunity thresholds [39] and in vaccination strategies. For example, the convenience of vaccinating network hubs first has been broadly discussed [40, 41], though that strategy is hampered by the difficulty of identifying actual hubs with local information. An interesting alternative consists in vaccinating neighbors of randomly chosen individuals, since these neighbors have more contacts on average [42]. Still, this latter strategy cannot consider protective behaviors exhibited by individuals, which might be independent of their contact habits. Perhaps an improved strategy should include at once a sensible use of this last strategy coupled with individual-dependent IFR, a quantity that depends not only on age, but also on sex, comorbidities and social roles, among others. Regardless the details of the model, the most efficient immunization strategy is conditioned by the availability of vaccines at each time, and optimization is particularly critical when availability is low-to-medium, while different strategies converge at sufficiently high vaccination rates.

We have focused in the reduction in the number of fatalities under different vaccination protocols, since this is a short-term benefit that can be easily quantified. However, different vaccination protocols also affect hospital occupation or the number of infections, and the variation can be very large depending on population structure, contact habits, and vaccination rate. The benefits of reducing the number of infections are more difficult to evaluate, but have to be kept in mind as our knowledge of the adaptive strategies of SARS-CoV-2 improves: more infections entail an enhanced number of circulating viral variants and therefore a higher probability of emergence of strains able to escape the protective effect of vaccines. In the mid and long term, it might be advisable to seek optimal vaccination strategies that simultaneously minimize the number of fatalities and the number of infections.

## Supporting information

Supplemental Material: Annexes 1 to 4

## Data Availability

All data produced in the present work are contained in the manuscript.

https://mybinder.org/v2/gh/IkerAtienza/SIYRD/main?urlpath=\%2Fvoila\%2Frender\%2FSimulator.ipynb

## Supporting information

**S1 Appendix. Calculation of contact matrices**

**S2 Appendix. Estimation of model parameters for COVID-19**

**S3 Appendix. Stability analysis of SIYRD fixed points**

**S4 Appendix. Application of S**^2^**IYRD to COVID-19, 50 years threshold**

## Acknowledgments

The authors are indebted to A. Vespignani for support with the use of their data. Grants PID2020-113284GB-C21 (I.A., S.M.) and BADS (PID2019-109320GB-100, S.A.) funded by MCIN/AEI/10.13039/501100011033. The Spanish MICINN has also funded the “Severo Ochoa” Centers of Excellence to CNB, SEV 2017-0712, and the special grant PIE 2020-20E079 (CNB) entitled “Development of protection strategies against SARS-CoV-2”.

## References

1. Atienza I. S^2^IYRD Model Simulator; 2021. https://mybinder.org/v2/gh/IkerAtienza/SIYRD/main?urlpath=%2Fvoila%2Frender%2FSimulator.ipynb.

2. Snowden FM. Epidemics and Society. Yale University Press. 2019. Available from: http://www.jstor.org/stable/j.ctvqc6gg5.

3. Rappuoli R, Pizza M, Del Giudice G, Gregorio E. Vaccines, new opportunities for a new society. Proceedings of the National Academy of Sciences of the United States of America. 2014;111. doi:10.1073/pnas.1402981111.

4. Antia A, Ahmed H, Handel A, Carlson NE, Amanna IJ, Antia R, et al. Heterogeneity and longevity of antibody memory to viruses and vaccines. PLOS Biology. 2018;16:1–15. doi:10.1371/journal.pbio.2006601.

5. Galanti M, Shaman J. Direct Observation of Repeated Infections With Endemic Coronaviruses. The Journal of Infectious Diseases. 2020;223(3):409–415. doi:10.1093/infdis/jiaa392.

6. Hansen CH, Michlmayr D, Gubbels SM, Mølbak K, Ethelberg S. Assessment of protection against reinfection with SARS-CoV-2 among 4 million PCR-tested individuals in Denmark in 2020: a population-level observational study. The Lancet. 2021;397(10280):1204–1212. doi:https://doi.org/10.1016/S0140-6736(21)00575-4.

7. Edridge AWD, Kaczorowska J, Hoste ACR, Bakker M, Klein M, Loens K, et al. Seasonal coronavirus protective immunity is short-lasting. Nature Medicine. 2020;26(11):1691–1693. doi:10.1038/s41591-020-1083-1.

8. Domingo-Calap P. Viral evolution and Immune responses. Journal of Clinical Microbiology and Biochemical Technology. 2019;5:013–018. doi:10.17352/jcmbt.000033.

9. Cobey S, Larremore D, Grad Y, Lipsitch M. Concerns about SARS-CoV-2 evolution should not hold back efforts to expand vaccination. Nature Reviews Immunology. 2021;21:1–6. doi:10.1038/s41577-021-00544-9.

10. Phillips N. The coronavirus will become endemic. Nature. 2021;590:382–384.

11. Lavine J, Bjornstad O, Antia R. Immunological characteristics govern the transition of COVID-19 to endemicity. Science. 2021;371:eabe6522. doi:10.1126/science.abe6522.

12. Bärnighausen T, Bloom DE, Cafiero-Fonseca ET, O’Brien JC. Valuing vaccination. Proceedings of the National Academy of Sciences. 2014;111(34):12313–12319. doi:10.1073/pnas.1400475111.

13. Rodrigues CMC, Plotkin SA. Impact of Vaccines; Health, Economic and Social Perspectives. Frontiers in Microbiology. 2020;11:1526. doi:10.3389/fmicb.2020.01526.

14. Bubar KM, Reinholt K, Kissler SM, Lipsitch M, Cobey S, Grad YH, et al. Model-informed COVID-19 vaccine prioritization strategies by age and serostatus. Science. 2021;371(6532):916–921. doi:10.1126/science.abe6959.

15. Rodríguez J, Patón M, Acuña JM. COVID-19 vaccination rate and protection attitudes can determine the best prioritisation strategy to reduce fatalities. medRxiv. 2021;doi:10.1101/2020.10.12.20211094.

16. Fitzpatrick MC, Galvani AP. Optimizing age-specific vaccination. Science. 2021;371(6532):890–891. doi:10.1126/science.abg2334.

17. Medlock J, Galvani AP. Optimizing Influenza Vaccine Distribution. Science. 2009;325(5948):1705–1708. doi:10.1126/science.1175570.

18. Mistry D, Litvinova M y, Piontti AP, Chinazzi M, Fumanelli L, Gomes MFC, et al. Inferring high-resolution human mixing patterns for disease modeling. Nature Communications. 2021;12(1). doi:10.1038/s41467-020-20544-y.

19. Prem K, Zandvoort Kv, Klepac P, Eggo RM, Davies NG, for the Mathematical Modelling of Infectious Diseases COVID-19 Working Group C, et al. Projecting contact matrices in 177 geographical regions: An update and comparison with empirical data for the COVID-19 era. PLOS Computational Biology. 2021;17:1–19. doi:10.1101/2020.07.22.20159772.

20. Makhoul M, Chemaitelly H, Ayoub HH, Seedat S, Abu-Raddad LJ. Epidemiological Differences in the Impact of COVID-19 Vaccination in the United States and China. Vaccines. 2021;9(3). doi:10.3390/vaccines9030223.

21. Goldstein E, Lipsitch M, Cevik M. On the Effect of Age on the Transmission of SARS-CoV-2 in Households, Schools, and the Community. The Journal of Infectious Diseases. 2020;223(3):362–369. doi:10.1093/infdis/jiaa691.

22. Davies NG, Klepac P, Liu Y, Prem K, Jit M, et al. Age-dependent effects in the transmission and control of COVID-19 epidemics. Nature Medicine. 2020;26(8):1205–1211. doi:10.1038/s41591-020-0962-9.

23. Zhang J, Litvinova M, Liang Y, Wang Y, Wang W, Zhao S, et al. Changes in contact patterns shape the dynamics of the COVID-19 outbreak in China. Science. 2020;368(6498):1481–1486. doi:10.1126/science.abb8001.

24. Mueller AL, McNamara MS, Sinclair DA. Why does COVID-19 disproportionately affect older people? Aging. 2020;12(10):9959–9981. doi:10.18632/aging.103344.

25. Liu Y, Mao B, Liang S, Yang JW, Lu HW, Chai YH, et al. Association between age and clinical characteristics and outcomes of COVID-19. European Respiratory Journal. 2020;55(5):2001112. doi:10.1183/13993003.01112-2020.

26. Finelli L, Gupta V, Petigara T, Yu K, Bauer KA, Puzniak LA. Mortality Among US Patients Hospitalized With SARS-CoV-2 Infection in 2020. JAMA Network Open. 2021;4(4):e216556. doi:10.1001/jamanetworkopen.2021.6556.

27. Pastor-Barriuso R, Pérez-Gómez B, Hernán MA, Pérez-Olmeda M, Yotti R, Oteo-Iglesias J, et al. Infection fatality risk for SARS-CoV-2 in community dwelling population of Spain: nationwide seroepidemiological study. BMJ. 2020; p. m4509. doi:10.1136/bmj.m4509.

28. To KKW, Hung IFN, Ip JD, Chu AWH, Chan WM, Tam AR, et al. Coronavirus Disease 2019 (COVID-19) Re-infection by a Phylogenetically Distinct Severe Acute Respiratory Syndrome Coronavirus 2 Strain Confirmed by Whole Genome Sequencing. Clinical Infectious Diseases. 2020;doi:10.1093/cid/ciaa1275.

29. Gupta V, Bhoyar RC, Jain A, Srivastava S, Upadhayay R, Imran M, et al. Asymptomatic Reinfection in 2 Healthcare Workers From India With Genetically Distinct Severe Acute Respiratory Syndrome Coronavirus 2. Clinical Infectious Diseases. 2020;doi:10.1093/cid/ciaa1451.

30. Tillett RL, Sevinsky JR, Hartley PD, Kerwin H, Crawford N, Gorzalski A, et al. Genomic evidence for reinfection with SARS-CoV-2: a case study. The Lancet Infectious Diseases. 2021;21(1):52–58. doi:10.1016/s1473-3099(20)30764-7.

31. Spiegelhalter D. Use of “normal” risk to improve understanding of dangers of COVID-19. BMJ. 2020;370. doi:10.1136/bmj.m3259.

32. Ilin C, Annan-Phan S, Tai X, Mehra S, Hsiang S, Blumenstock J. Public Mobility Data Enables COVID-19 Forecasting and Management at Local and Global Scales. Scientific Reports. 2021;11:13531. doi:10.1101/2020.10.29.20222547.

33. Wellenius GA, Vispute S, Espinosa V, Fabrikant A, Tsai TC, Hennessy J, et al. Impacts of social distancing policies on mobility and COVID-19 case growth in the US. Nature Communications. 2021;12(1). doi:10.1038/s41467-021-23404-5.

34. Weycker D, Edelsberg J, Halloran ME, Longini IM, Nizam A, Ciuryla V, et al. Population-wide benefits of routine vaccination of children against influenza. Vaccine. 2005;23(10):1284–1293. doi:10.1016/j.vaccine.2004.08.044.

35. Rodríguez J, Patón M, Acuña JM. COVID-19 vaccination rate and protection attitudes can determine the best prioritisation strategy to reduce fatalities. medRxiv. 2020;doi:10.1101/2020.10.12.20211094.

36. Han S, Cai J, Yang J, Zhang J, Wu Q, Zheng W, et al. Time-varying optimization of COVID-19 vaccine prioritization in the context of limited vaccination capacity. Nature Communications. 2021;12. doi:10.1038/s41467-021-24872-5.

37. Moore S, Hill EM, Dyson L, Tildesley MJ, Keeling MJ. Modelling optimal vaccination strategy for SARS-CoV-2 in the UK. PLOS Computational Biology. 2021;17(5):1–20. doi:10.1371/journal.pcbi.1008849.

38. Matrajt L, Eaton J, Leung T, Brown E. Vaccine optimization for COVID-19, who to vaccinate first? Science Advances. 2021;7. doi:10.1101/2020.08.14.20175257.

39. Pastor-Satorras R, Vespignani A. Epidemic Spreading in Scale-Free Networks. Physical review letters. 2001;86:3200–3. doi:10.1103/PhysRevLett.86.3200.

40. Pastor-Satorras R, Vespignani A. Immunization of Complex Networks. Physical review E, Statistical, nonlinear, and soft matter physics. 2002;65:036104. doi:10.1103/PhysRevE.65.036104.

41. Dezso Z, Barabasi AL. Halting viruses in scale-free networks. Physical review E, Statistical, nonlinear, and soft matter physics. 2002;65:055103. doi:10.1103/PhysRevE.65.055103.

42. Cohen R, Havlin S, ben Avraham D. Efficient Immunization Strategies for Computer Networks and Populations. Physical review letters. 2004;91:247901. doi:10.1103/PhysRevLett.91.247901.

