## Supplemental Material: Annexes 1 to 4 for "Vaccination strategies in structured populations under partial immunity and reinfection"

### Supplementary file 1: **Calculation of contact matrices**

The contact matrix plays a critical role in the $S^2$IYRD model. We have used the available contact matrices derived in [1]. They are $85 \times 85$ matrices whose elements, $M_{ij}$, represent the number of contacts an individual of age $i$ has with individuals of age $j$. Each row and column refers to a single year of age —starting at age 0—, except for the last row and column, which contain ages 84 and older.

#### Definition of the contact matrix

For each of the four countries analyzed in the main text, we additionally updated its corresponding contact matrix by incorporating the most recent demographic information (see S2 Appendix). To do so, we revisit here how each contact matrix is originally calculated (see [1] for more information). Contact matrices distinguish several different places where contacts occur, also known as settings: household (H), school (S), workplace (W) and general community (GC). First, the relative abundance of contacts between individuals of age $i$ and individuals of age $j$ in each instance $s$ of the setting $k$, $\Gamma^{k(s)}$ (symmetrical matrices) was calculated from local and national surveys. For each setting $k$, the age-based contact patterns are encoded in a non-symmetrical contact matrix $F^k$, whose elements $F^k_{ij}$ describe the average frequency of contact between a given individual of age $i$ and individuals of age $j$ in setting $k$,

$$F^k_{ij} = \sum_{s:\nu^{k(s)}>1} \Gamma^{k(s)}_{ij}/N_i \,, \tag{1}$$

where $N_i$ is the total number of individuals of age $i$. Since both $F^k$ and the original demographic data were available, we could easily update $F^k$ and, hence, the contact matrices, using the new demographic structure and Eq. (1). Finally, the contact matrix $M$ is obtained as the weighted linear combination of these four setting-associated contact matrices.

$$M_{ij} = \sum_k w_k F^k_{ij} \,, \tag{2}$$

where $w_k$ is the average number of contacts in each specific setting for all the individuals in the community previously estimated by [1]. Although matrix $\mathbf{M}$ is not symmetrical, it satisfies the closure relation

$$M_{ij} N_i = M_{ji} N_j \tag{3}$$

#### Transformation of the contact matrix

Besides a demographic update, contact matrices have to be transformed to properly represent population separation into groups. We summarize this through the following general procedure for an arbitrary number of groups, particularizing to the case of two final groups.

**Example: from a $4 \times 4$ to a $2 \times 2$ matrix**

As an example, let us start with a contact matrix $G$ with four different population groups that have to be grouped into two. Schematically,

$$G = \begin{pmatrix} G_{11} & G_{12} & G_{13} & G_{14} \\ G_{21} & G_{22} & G_{23} & G_{24} \\ G_{31} & G_{32} & G_{33} & G_{34} \\ G_{41} & G_{42} & G_{43} & G_{44} \end{pmatrix} \to H = \begin{pmatrix} H_{11} & H_{12} \\ H_{21} & H_{22} \end{pmatrix} ,$$

where $N_i$ represents the number of individuals in each group and the matrix $G$ satisfies the closure relation (3). Assume also that group stratification is as follows,

$$G_A = \{G_1, G_2\} \qquad G_B = \{G_3, G_4\} \tag{4}$$

By definition, $H_{11}$ is the number of contacts an individual from $G_A$ has with individuals from that same group. Since $G_A$ is formed by two groups, we need to account for two different contributions: intra ($G_1 \leftrightarrow G_1$ and $G_2 \leftrightarrow G_2$) and inter-group contacts ($G_1 \leftrightarrow G_2$). Hence, if each $G_1$-individual has $G_{11}$ contacts with other $G_1$-individuals, the total number of $G_1$-intra contacts will be $G_{11}N_1$. In the case of $G_2$-intra contacts, the total number will be $G_{22}N_2$. Regarding the total number of inter-group contacts, due to the closure relation, we can consider them from a $G_1$-individual perspective ($G_{12}N_1$) or from a $G_2$-individual perspective ($G_{21}N_2$). We can also account for both contributions and divide them by 2. Finally, if we add inter- and intra contributions and divide by the number of individuals in $G_A$, we obtain

$$H_{11} = \frac{G_{11}N_1 + G_{22}N_2 + \frac{1}{2}(G_{12}N_1 + G_{21}N_2)}{N_1 + N_2} \tag{5}$$

and, similarly,

$$H_{22} = \frac{G_{33}N_3 + G_{44}N_4 + \frac{1}{2}(G_{34}N_3 + G_{43}N_4)}{N_3 + N_4} \tag{6}$$

Conversely, $H_{12}$ is the number of contacts a $G_A$-individual has with $G_B$-individuals. As before, this can be decomposed into subgroups. The total number of $G_B$-contacts a $G_1$-individual has is $(G_{13} + G_{14})N_1$; similarly, a $G_2$-individual has $(G_{23} + G_{24})N_2$ $G_B$ contacts. By summing both contributions and dividing them by the total number of individuals $G_A$ we arrive to:

$$H_{12} = \frac{(G_{13} + G_{14})N_1 + (G_{23} + G_{24})N_2}{N_1 + N_2} , \tag{7}$$

$$H_{21} = \frac{(G_{31} + G_{31})N_3 + (G_{41} + G_{42})N_4}{N_3 + N_4} \tag{8}$$

As it can be checked, the projected matrix $H$ also satisfies the closure relation, $H_{ij}N_i = H_{ji}N_j$.

##### General transformation into two groups

Let us now derive the general expression of the 2x2 matrix starting from an $n \times n$ matrix,

$$G = \begin{pmatrix} G_{11} & G_{12} & \dots & G_{1n} \\ G_{21} & G_{22} & \dots & \\ \vdots & & \ddots & \\ G_{n1} & & & G_{nn} \end{pmatrix} \rightarrow H = \begin{pmatrix} H_{11} & H_{12} \\ H_{21} & H_{22} \end{pmatrix}$$

Let us set the threshold between the two final groups ($H_1$ and $H_2$) at sub-group $x$ in such a way that

$$H_1 = \bigcup_{i=1}^{x} G_i \qquad H_2 = \bigcup_{i=x+1}^{n} G_i \tag{9}$$

where, as previously, $N_i$ is the number of individuals in group $G_i$. $H_{12}$ and $H_{21}$ are easily obtained by extending (7) and (8) to an $n \times n$ matrix. $H_{12}$ is the number of contacts an $H_1$-individual has with $H_2$-individuals. Therefore, we must compute first the total number of contacts each sub-group contained in $H_1$ has with every sub-group in $H_2$ ($N_i \sum_{j=x+1}^{n} G_{i,j}$), repeat for all subgroups in $H_1$, and add the resulting terms. This corresponds to the numerator of the first expression in (10), while the denominator is just the sum of all individuals within the $H_1$ group. The calculation proceeds analogously for group $H_2$, yielding:

$$H_{12} = \frac{\sum_{i=1}^{x} N_i \left( \sum_{j=x+1}^{n} G_{ij} \right)}{\sum_{i=1}^{x} N_i}, \quad H_{21} = \frac{\sum_{j=x+1}^{n} N_j \left( \sum_{i=1}^{x} G_{ji} \right)}{\sum_{j=x+1}^{n} N_j} \tag{10}$$

$$H_{11} = \frac{\sum_{i=1}^{x} G_{ii} N_i + \frac{1}{2} \sum_{i=1}^{x} N_i \left( \sum_{\substack{j=1 \\ j \neq i}}^{x} G_{ij} \right)}{\sum_{i=1}^{x} N_i} \tag{11}$$

$$H_{22} = \frac{\sum_{j=x+1}^{n} G_{jj} N_j + \frac{1}{2} \sum_{j=x+1}^{n} N_j \left( \sum_{\substack{i=x+1 \\ i \neq j}}^{n} G_{ji} \right)}{\sum_{j=x+1}^{n} N_j} \tag{12}$$

In the case of the diagonal entries, Eqs. (11) and (12), we get two different summation terms in the numerator. The first sum in Eq. (11) corresponds to the first two terms appearing in Eq. (5). In this way, $G_{ii} N_i$ represents the total number of contacts of individuals within the same subgroup $i$ ($G_i \in H_1$). The summation collects all $H_1$ contributions.

The second term in Eq. (11) is the equivalent to the last term in the numerator of Eq. (5). The effect of merging different subgroups within $H_1$ is that now the entries in the main diagonal of $H$ must incorporate not only the contacts between individuals within the same subgroup, but also the contacts between individuals of different $H_1$ sub-groups. The total number of contacts between $G_i$ individuals and the remaining sub-groups in $H_1$ is given by this second sum in Eq. (11). The factor $\frac{1}{2}$ corrects the double counting of these inter sub-group contacts. Eqs. (11) and (12) can be expressed in a more compact way rearranging some terms:

$$H_{11} = \frac{\sum_{i=1}^{x} N_i \left[ G_{ii} + \sum_{j=1}^{x} G_{ij} \right]}{2 \sum_{i=1}^{x} N_i} \tag{13}$$

$$H_{22} = \frac{\sum_{j=x+1}^{n} N_j \left[G_{jj} + \sum_{i=x+1}^{n} G_{ji}\right]}{2\sum_{j=x+1}^{n} N_j} \tag{14}$$

**Transformation into an arbitrary number of groups**

To conclude this section, hereafter we take the generalization a step further by explaining how to transform an $n \times n$ contact matrix into a matrix with $m$ population groups,

$$G = \begin{pmatrix} G_{11} & G_{12} & \dots & G_{1n} \\ G_{21} & G_{22} & \dots & \\ \vdots & & \ddots & \\ G_{n1} & & & G_{nn} \end{pmatrix} \rightarrow H = \begin{pmatrix} H_{11} & H_{12} & \dots & H_{1m} \\ H_{21} & H_{22} & \dots & \\ \vdots & & \ddots & \\ H_{m1} & & & H_{mm} \end{pmatrix}$$

Now, $m_1, m_2, ..., m_{m-1}$ represent the limits between the age groups (with $m_m = n$ and $m_0 = 0$). Again, $G_{ij}$ represents the number of contacts an individual of age $i$ has with individuals of age $j$ and $N_i$ is the number of individuals of age $i$.

Generalizing the derivations above, entries of matrix $\mathbf{H}$ are obtained from

$$H_{ii} = \frac{\sum_{k=m_{(i-1)}+1}^{m_i} G_{kk} N_k + \frac{1}{2}\left[\sum_{k=m_{(i-1)}+1}^{m_i} N_k \left(\sum_{\substack{l=m_{(i-1)}+1 \\ l \neq k}}^{m_i} G_{kl}\right)\right]}{r\sum_{k=m_{(i-1)}+1}^{m_i} N_k} \tag{15}$$

$$H_{ij} = \frac{\sum_{k=m_{(i-1)}+1}^{m_i} N_k \left(\sum_{l=m_{(j-1)}+1}^{m_j} G_{kl}\right)}{\sum_{k=m_{(i-1)}+1}^{m_i} N_k} \tag{16}$$

Where $1 \leq i, j \leq m$ and $m_0 = 0$, $m_m = n$. Finally, Eq. (15) can be written as

$$H_{ii} = \frac{\sum_{k=m_{(i-1)}+1}^{m_i} N_k \left[G_{kk} + \sum_{l=m_{(i-1)}+1}^{m_i} G_{kl}\right]}{2\sum_{k=m_{(i-1)}+1}^{m_i} N_k}. \tag{17}$$

### References


1. Mistry D, Litvinova M, y Piontti AP, Chinazzi M, Fumanelli L, Gomes MFC, et al. Inferring high-resolution human mixing patterns for disease modeling. Nature Communications. 2021;12(1). doi:10.1038/s41467-020-20544-y.

Supplementary file 2: **Estimation of model parameters for COVID-19**

#### Demographic data

During the evolution of the COVID-19 pandemic, the SARS-CoV-2 virus has impinged differently on different age groups. Therefore, information on the demographic structure of a population should be included in simulations of its dynamics as a possible main factor affecting it.

Figure 1 displays demographic profiles for the four countries studied in the main text. They have been chosen to highlight the high variability of demographic pyramids. In these cases, both the shape of the demography and the size of the populations ($N$) vary greatly: Israel has $8,655,540$ inhabitants, Spain $47,332,613$, the South African Republic (SAR) $59,308,689$, and the United States of America (USA) $331,002,647$. In the case of Spain, census data from the National Institute of Statistics [1] are used, considering the figures as of January 1, 2020. The demographic information for the rest of the countries has been obtained from the UN World Population Prospects 2019 [2].

In the S$^2$IYRD model, the original population is divided into two groups, $G_1$ and $G_2$, with $N_1$ and $N_2$ individuals, respectively, by selecting a specific age threshold. All variables depend on the country of choice. A main variable we consider is the fraction of population in the older group, $n_2 = N_2/N$, (by definition, $n_1 = 1 - n_2$). Figure 2 shows the fraction of individuals in group $G_2$ for two different age thresholds and the four countries in Fig. 1 (note the logarithm in the $y-$axis); see also S4 Appendix.

We have included three additional countries on the website [3] that are not analyzed in depth in the main text. The demographic information for India, Italy and Japan corresponds to 2020 data [2].

#### Infection fatality risk estimates

The infection fatality risk (IFR) of a contagious disease is a measure of disease severity. The IFR is defined as the ratio between the number of deaths caused by the disease and the total number of infections in a given time interval. Formally, it is estimated as

$$IFR_i = \frac{Deaths_i}{Infected_i} \tag{1}$$

where $i$ specifies each of the groups considered. The data needed to calculate the IFR, however, are not easy to obtain and classify. In the context of the ongoing COVID-19 pandemic, the lack of sufficient testing, especially in the first months of the pandemic, prevents an accurate estimation of the total number of infections and deaths, a problem compounded by the unknown number of asymptomatic infections caused by SARS-CoV-2.

Beyond depending on the disease under consideration, the fatality risk varies with characteristics of individuals such as age, sex, ethnic group, or comorbidities. For this reason, we independently calculate the estimation of the IFR of COVID-19 for each of the populations represented in Fig. 1. Data for each country was obtained from their own national sources. In the case of Spain, we considered the updated official data provided by the Carlos III National Institute of Health as of March 8, 2021 [4]. For

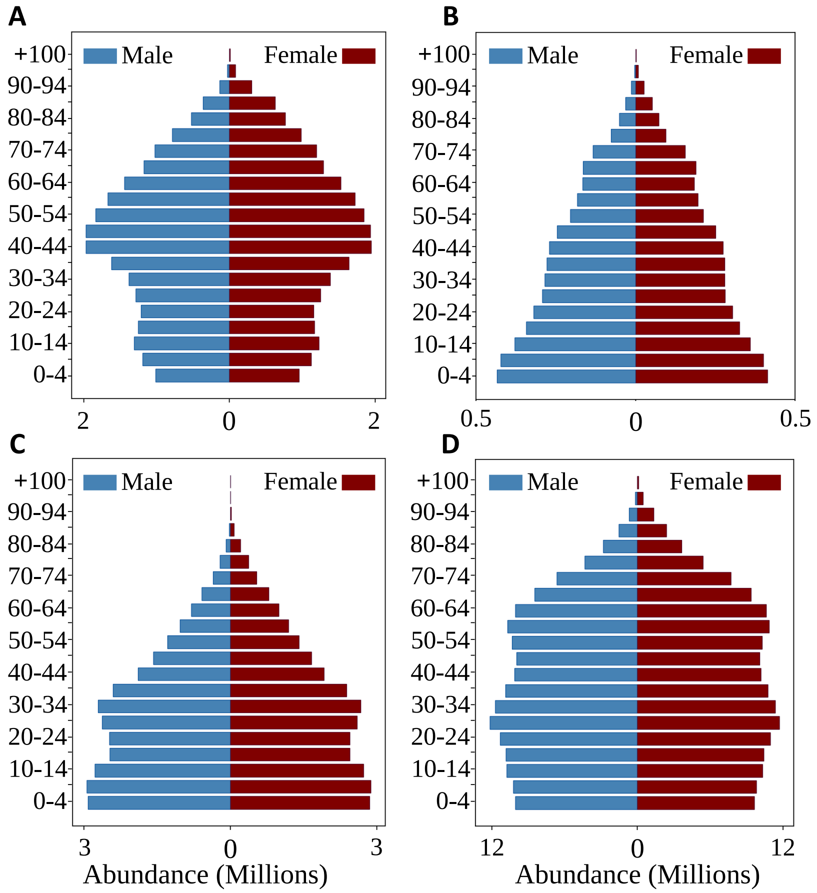


**Fig 1.** Demographic pyramid for the (A) Spanish (B) Israeli (C) South African and (D) U.S. population in 2020; in the vertical axis, populations are grouped in 5-year intervals, starting with 0-4 years at the bottom. The last group includes people aged 100 and older.

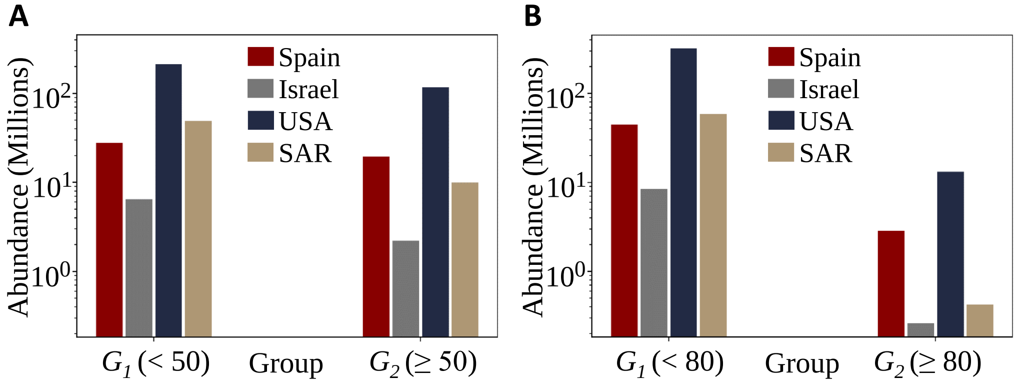


**Fig 2.** Absolute sizes $N_1$ and $N_2$ of groups $G_1$ and $G_2$ for two different age thresholds, (A) 50 and (B) 80 years. In the vertical axis, numbers represents millions of people in log-scale. Relative group sizes $n_1$ and $n_2$ are reported in S4 Appendix.

Israel, the number of cases was obtained directly from government reports [5], while the number of deaths was downloaded from the Israel Science and Technology Directory [6] as of June 10, 2021. Data for the South African Republic were obtained from the National Institute For Communicable Diseases [7] [8] as of June 26, 2021 (Week 25 reports). Finally, U.S. data were provided by the Center for Disease Control and Prevention [9], updated as of August 4, 2021.

Figure 3 summarizes the IFR of the Spanish, Israeli, South African an U.S. population in 10-years age groups. The exponential growth of the IFR with age for all four countries analyzed is clear and comparable, despite some variability at the younger ages (whose data is, however, less reliable from a statistical viewpoint).

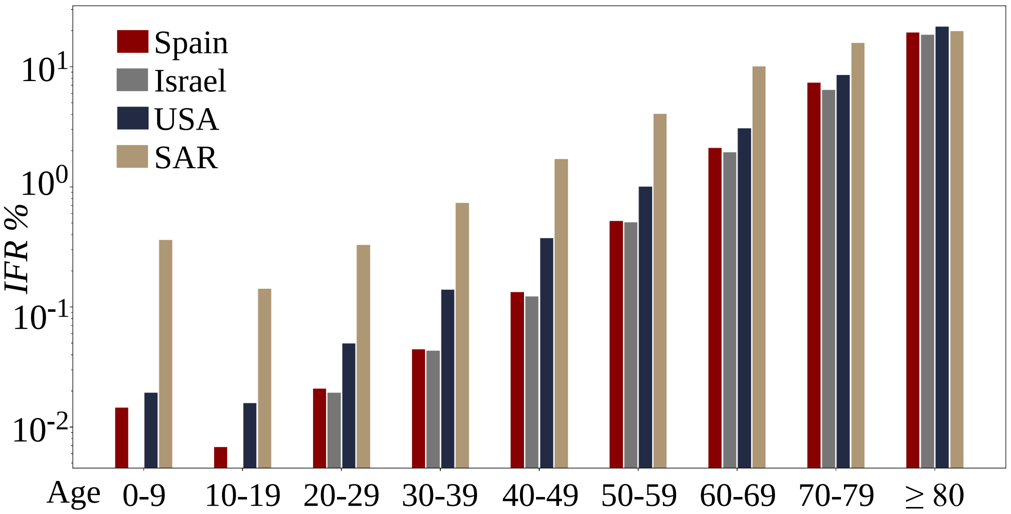


**Fig 3.** COVID-19 Infection Fatality Risk by 10-year age groups; in the vertical axis, numbers represent IFR percentages in log-scale (only every second label is displayed).

The heterogeneity of these nine groups is projected in only two groups in the framework of our model. To do so, we recalculate the IFR by summing up the weighted contributions of groups below ($IFR_1$) and above ($IFR_2$) the considered age threshold. Figure 4 shows results for two different age thresholds.

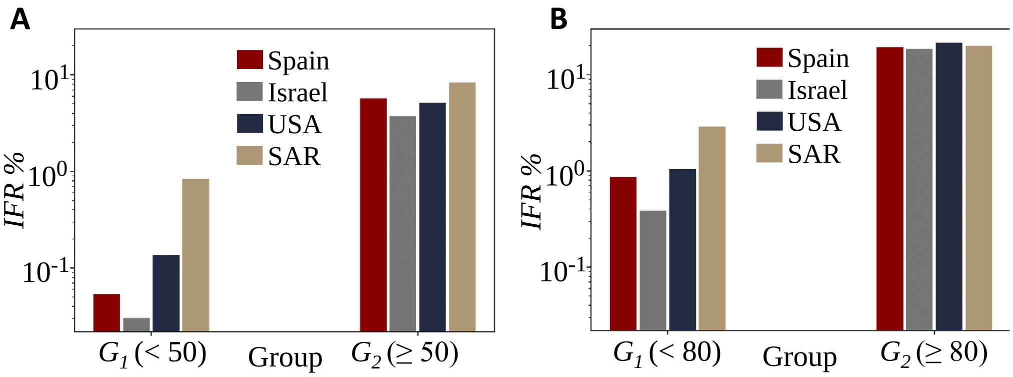


**Fig 4.** COVID-19 Infection Fatality Risk by group for two different age thresholds, 50 (A) and 80 (B) years; in the vertical axis, numbers represent IFR percentages in log-scale.

Differences between countries in $IFR_1$ are remarkable, while $IFR_2$ values are more similar to each other, regardless the age threshold considered. Table 1 reports the ratio between the two IFR in calculated, quantifying the highly different impact that COVID-19 has as a funcion of age.

**Table 1.** Empirical IFR ratio ($IFR_2/IFR_1$) for the countries in Figure 1.

| **IFR ratio, $\mathbf{IFR_2/IFR_1}$** | | | | |
|---|---|---|---|---|
| Age limit | Spain | Israel | SAR | USA |
| 50 years | 105.99 | 123.14 | 9.89 | 37.79 |
| 80 years | 22.18 | 48.19 | 6.90 | 20.63 |

As above, data on cases and deaths for those additional countries included in the website were obtained from their own national sources —with the exception of India, where we used the Spanish IFR because we could not find the corresponding data. In the case of Italy, deaths and cases were provided by the Italian National Institute of Health [10] as of June 23, 2021. Japanese data were provided by the Ministry of Health, Labor and Welfare [11] as of June 30, 2021.

#### Infection and death rates

For all simulations performed, infection rates were computed as:

$$\beta_{SI} = \frac{R_0}{d_I} \qquad \beta_{RI} = \alpha_1 \beta_{SI} \qquad \beta_{RY} = \alpha_2 \beta_{RI} \qquad \beta_{SY} = \frac{\beta_{RY} \cdot \beta_{SI}}{\beta_{RI}} \tag{2}$$

where $R_0$ is the reproductive number of the epidemic disease under consideration, $d_I$ is the infectious period (the time interval during which the individual is infectious), and $\alpha_i$ are two constants satisfying $\alpha_i \leq 1$. Since our model lacks an exposed compartment (E), we subsume under $d_I$ the average exposure period of infected individuals, estimated at 3 days [12]. Furthermore, available data suggest that patients with mild to moderate COVID-19 remain infectious no longer than 10 days after symptom onset [13]: therefore, we take $d_I = 13$ days.

The recovery and mortality rates of primary infections are functions of the infection fatality risk (IFR) and the infectious period ($d_I$),

$$r_i = \frac{1 - IFR_i}{d_I} \tag{3}$$

$$\mu_{I_i} = \frac{IFR_i}{d_I}, \tag{4}$$

where sub-indexes correspond to each age group.

The vaccination rate is a constant percentage of the total population ($N$). In the age-stratified model $\mathrm{S^2IYRD}$, it was assumed that at most 70% of the individuals in each age group could be vaccinated. Group prioritization strategies limited vaccination to one age group at a time while, for the simultaneous vaccination strategy, the total vaccination rate is multiplied by the relative group abundance of each group, $n_i/N$. When the vaccination threshold of 70% is reached, vaccination starts in the other group, provided the number of susceptible individuals at that time is still larger than $0.3n_i$; otherwise, vaccination halts.

### References


1. Instituto Nacional de Estadística. Resident population by date, sex and age. Last accessed: August 18, 2021;. https://www.ine.es/jaxiT3/Tabla.htm?t=31304.

2. Population Pyramid. Population Pyramids of the World from 1950 to 2100. Last accessed: August 18, 2021; 2019. https://www.populationpyramid.net/world/2020.

3. Atienza I. S$^2$IYRD Model Simulator; 2021. https://mybinder.org/v2/gh/IkerAtienza/SIYRD/main?urlpath=%2Fvoila%2Frender%2FSimulator.ipynb.

4. Instituto de Salud Carlos III. Datos oficiales de COVID-19 en España. Last accessed: March 09, 2021; 2021. https://github.com/rubenfcasal/COVID-19/tree/master/historico_csv/21_03_09.

5. DataGov. Corona data by sex and age groups. Last accessed: June 10, 2021; 2021. https://data.gov.il/dataset/covid-19/resource/89f61e3a-4866-4bbf-bcc1-9734e5fee58e.

6. Israel Science and Technology Directory. The age distribution and sex of the patients who died with coronavirus (COVID-19) infection. Last accessed: June 10, 2021; 2021. https://www.science.co.il/medical/coronavirus/Death-statistics.php.

7. National Institute For Communicable Diseases. Weekly hospital surveillance (DATCOV) update. Last accessed: June 30, 2021; 2021. https://www.nicd.ac.za/diseases-a-z-index/disease-index-covid-19/surveillance-reports/weekly-hospital-surveillance-datcov-update/.

8. National Institute For Communicable Diseases. COVID-19 weekly epidemiology brief (Week 25 2021). Last accessed: June 30, 2021; 2021. https://www.nicd.ac.za/wp-content/uploads/2021/07/COVID-19-Weekly-Epidemiology-Brief-week-25-2021.pdf.

9. Centers for Disease Control and Prevention. COVID-19 Case Surveillance Public Use Data. Last accessed: August 04, 2021; 2021. https://data.cdc.gov/Case-Surveillance/COVID-19-Case-Surveillance-Public-Use-Data/vbim-akqf.

10. Istituto Superiore di Sanità. COVID-19 Epidemic National Update. Last accessed: June 23, 2021; 2021. https://www.epicentro.iss.it/coronavirus/bollettino/Bollettino-sorveglianza-integrata-COVID-19_23-giugno-2021.pdf.

11. Toyo Keizai Online COVID-19 Task Team. Coronavirus Disease (COVID-19) Situation Report in Japan. Last accessed: June 30, 2021; 2020. https://toyokeizai.net/sp/visual/tko/covid19/en.html.

12. Bubar KM, Reinholt K, Kissler SM, Lipsitch M, Cobey S, Grad YH, et al. Model-informed COVID-19 vaccine prioritization strategies by age and serostatus. Science. 2021;371(6532):916–921. doi:10.1126/science.abe6959.

13. Centers for Disease Control and Prevention. Ending Isolation and Precautions for People with COVID-19: Interim Guidance. Last Accesed: March 26, 2021; 2021. https://www.cdc.gov/coronavirus/2019-ncov/hcp/duration-isolation.html.

### Supplementary file 3: **Stability analysis of SIYRD fixed points**

##### Fixed points

Considering $n = S + I + Y + R + D = 1$ and excluding the redundant equation for the number of deaths, SIYRD equations are simplified to

$$\dot{S} = -\beta_{SI} IS - \beta_{SY} YS - v\Theta(S) \quad (1)$$

$$\dot{I} = \beta_{SI} IS + \beta_{SY} YS - rI - \mu_I I \quad (2)$$

$$\dot{Y} = \beta_{RI} IR + \beta_{RY} YR - rY - \mu_Y Y \quad (3)$$

$$\dot{R} = rI + rY - \beta_{RI} IR - \beta_{RY} YR + v\Theta(S) \quad (4)$$

For convenience, we will now take a explicit form for $\Theta(S)$, such that $\Theta(S) \to 1$ when $S \ll n$ and $\Theta(S) \to 0$ when $S \to (1-\theta)$, with $\theta$ the population fraction that can be vaccinated. In our simulations, and in agreement with actual estimations, we fix $\theta = 0.7$. In the following, we assume a Hill function for $\Theta(S)$,

$$\Theta(S) = \frac{S^z}{S^z + k^z}\,, \quad (5)$$

with $z > 1$ being an integer.

Setting Eqs. (1-4) to zero yields the only positive steady state of the system:

$$X_1^* = (0, 0, 0, R^*, D^*) \qquad | \qquad R^* + D^* = n = 1 \quad (6)$$

The asymptotic value of $R^*$ (and therefore of $D^*$) can be computed under certain circumstances. Setting $I^* = 0$ and $S^* = 0$ in (3) and (4) yields two different values of $R^*$ satisfying each equation, respectively

$$(\beta_{RY} R - r - \mu_Y)Y = 0 \to R_1^* = \frac{r + \mu_Y}{\beta_{RY}} \quad (7)$$

$$(r - \beta_{RY} R)Y = 0 \to R_2^* = \frac{r}{\beta_{RY}}\,. \quad (8)$$

For initial conditions with no recovered individuals, only $R_2^*$ can be ever reached. This will occur if, and only if, the number of recovered individuals reaches that value before the number of reinfected individuals drops to zero,

$$R^* = R_2^* = \frac{r}{\beta_{RY}} \qquad \text{if} \begin{cases} Y(t = t_1) = 0 \\ R(t = t_0) = \dfrac{r}{\beta_{RY}} \end{cases} \qquad \text{with } t_0 < t_1\,. \quad (9)$$

In the opposite situation, for $Y(t) = 0$ and $R(t) < R_2^*$, the steady-state solution cannot be obtained in explicit analytical form.

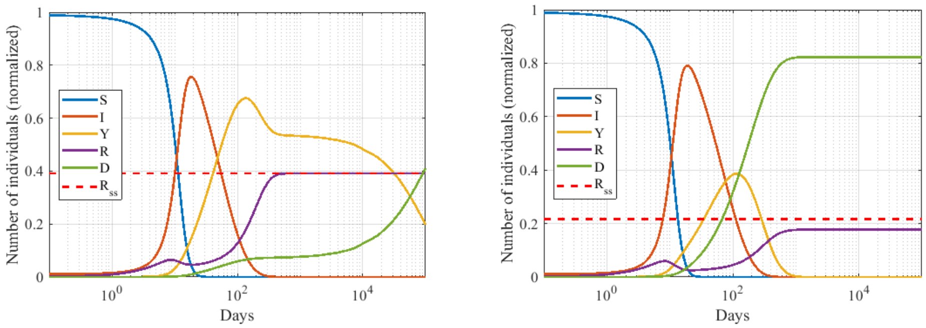


**Fig 1.** Generic dynamics of the SIYRD model. Comparison of analytical (left) and non-analytical (right) steady-state expression (note the logarithmic $x-$axis). Parameters are $\beta_{SI} = \beta_{RI} = 0.46$, $\beta_{SY} = \beta_{RY} = 0.046$, $\mu_I = 0.00149$ (left), $\mu_Y = 10^{-5}$ (left), $\mu_I = \mu_Y = 0.0045$ (right), $r = 0.0180$ (left), $r = 0.010$ (right), $v = 0.01$, $\theta = 0.7$ and Hill coefficient $z = 8$. The red dashed line represents $R_{ss} = R_2^*$. On the left, $R^* = R_{ss}$ exemplifying a case where the asymptotic value can be analytically calculated; on the right, $R^* \neq R_{ss}$ prevents from obtaining an analytical expression of the steady state. Initial conditions: $(0.99, 0.01, 0, 0, 0)$.

#### Linear stability analysis for $v \neq 0$

The linear stability of $X_1^*$ is given by the eigenvalues of its associated Jacobian matrix. The Jacobian of the SIYRD model, Eqs. (1-4) evaluated at $X_1^*$ is

$$J(X_1^*) = \begin{pmatrix} 0 & 0 & 0 & 0 \\ 0 & -r-\mu_I & 0 & 0 \\ 0 & \beta_{RI}R^* & \beta_{RY}R^* - r - \mu_Y & 0 \\ 0 & r - \beta_{RI}R^* & r - \beta_{RY}R^* & 0 \end{pmatrix}, \tag{10}$$

Since $J(X_1^*)$ is a lower triangular matrix, the eigenvalues correspond to its diagonal entries,

$$\lambda_1 = 0 \qquad \lambda_2 = -r - \mu_I \qquad \lambda_3 = \beta_{RY}R^* - r - \mu_Y \qquad \lambda_4 = 0 \tag{11}$$

Since there are two null eigenvalues and $\lambda_2$ is negative ($\lambda_2 < 0\, \forall r$, $\mu_Y > 0$), the stability condition reads $\lambda_3 \leq 0$, that is, $R^* \leq (r + \mu_Y/\beta_{RY})$. This inequality always holds because $R^* \leq R_2^*$ always, as discussed before. Therefore, the steady state $X_1^*$ is neutrally stable.

The previous stability analysis can be easily extended to the situation in which there is no infection of recovered individuals by reinfected ones ($\beta_{RY} = 0$). The only difference is that now, once $I = S = 0$, $R$ increases monotonically until $Y$ reaches zero. This means that equations (3) and (4) decouple and, therefore, there is no general analytical expression for the value of $R$ that satisfies their equilibria simultaneously. In spite of this modification, the Jacobian of the system is just $J(X_1^*; \beta_{RY} = 0)$ and the stability condition mentioned before directly disappears, resulting again in the same neutrally stable fixed point.

##### Linear stability analysis for $v = 0$

In the absence of vaccination, the system displays two different steady states. The first one is again $X_1^*$, and it remains always neutrally stable. The new steady state is:

$$X_2^* = (S^*, 0, 0, R^*, D^*) \tag{12}$$

$$J(X_2^*) = \begin{pmatrix} 0 & -\beta_{SI}S^* & -\beta_{SY}S^* & 0 \\ 0 & \beta_{SI}S^* - r - \mu_I & \beta_{SY}S^* & 0 \\ 0 & \beta_{RI}R^* & \beta_{RY}R^* - r - \mu_Y & 0 \\ 0 & r - \beta_{RI}R^* & r - \beta_{RY}R^* & 0 \end{pmatrix}$$

The Jacobian at $X_2^*$ yields a matrix which is not triangular. Still, its eigenvalues can be obtained with relative ease. Calculating the determinant $|J(X_2^*) - \lambda\mathbb{I}|$ by adjoints leads to the characteristic polynomial equation,

$$\lambda^2((A-\lambda)(D-\lambda) - BC) = \lambda^2(\lambda^2 - (A+D)\lambda + AD - BC) = 0, \tag{13}$$

where $A = \beta_{SI}S^* - (r + \mu_I)$, $B = \beta_{SY}S^*$, $C = \beta_{RI}R^*$ and $D = \beta_{RY}R^* - (r + \mu_Y)$.

Although $\lambda_1 = \lambda_2 = 0$, one needs to solve a quadratic equation to compute the other two eigenvalues. However, instead of doing that, it is possible to look at their signs to determine stability. Then, the determinant of the corresponding sub-matrix would be:

$$\Delta = AD - BC = (\beta_{SI}\beta_{RY} - \beta_{SY}\beta_{RI})R^*S^* - (r + \mu_Y)S^* - (r + \mu_I)R^* \tag{14}$$

Using $\frac{\beta_{SY}}{\beta_{SI}} = \frac{\beta_{RY}}{\beta_{RI}}$, Eq. (14) can be written as

$$\Delta = -(r + \mu_Y)S^* - (r + \mu_I)R^* < 0 \tag{15}$$

Therefore, $\Delta = \lambda_3\lambda_4 < 0$ meaning that $\lambda_3$ and $\lambda_4$ are both real with opposite sign. This implies that $X_2^*$ is an unstable fixed point. The only initial condition that leads to this steady state is, precisely, $X(t=0) = X_2^*$. This solution is irrelevant from an epidemiological perspective, since there is no epidemic in the absence of infected individuals that trigger it ($I(0) = Y(0) = 0$).

##### Linear stability analysis for $\mu_Y = 0$

If the mortality of reinfected individuals is set to zero ($\mu_Y = 0$), $X_1^*$ appears again, retaining its neutral stability. In addition, a new epidemic steady state emerges,

$$X_3^* = (0, 0, Y^*, R^*, D^*) \text{ with } R^* = \frac{r}{\beta_{RY}} \tag{16}$$

The characteristic equation $|J(X_3^*) - \lambda\mathbb{I}| = 0$ is now

$$(-\beta_{SY}Y^* - \lambda)(-r - \mu_I - \lambda)[\lambda^2 - (A+D)\lambda + AD - BC] = 0\,, \tag{17}$$

where $A = \beta_{RY}R^* - r$ , $B = \beta_{RY}Y^*$ , $C = r - \beta_{RY}R^*$ and $D = -\beta_{RY}Y^*$. Since $AD - BC = 0$ (replacing $R^*$ using (16)), the last two eigenvalues can be directly obtained: $\lambda_3 = 0$ and $\lambda_4 = -Y^*$. Hence, $X_3^*$ is another neutrally stable fixed point. As it occurred in the general case, the question is whether the system reaches first $R(t) = R^*$ ($X_3^*$ is the steady state) or $Y(t) = 0$ ($X_1^*$ steady state). Notice also that despite knowing the asymptotic value of $R$, $R^*$, if $X_3^*$ is the steady state reached by the

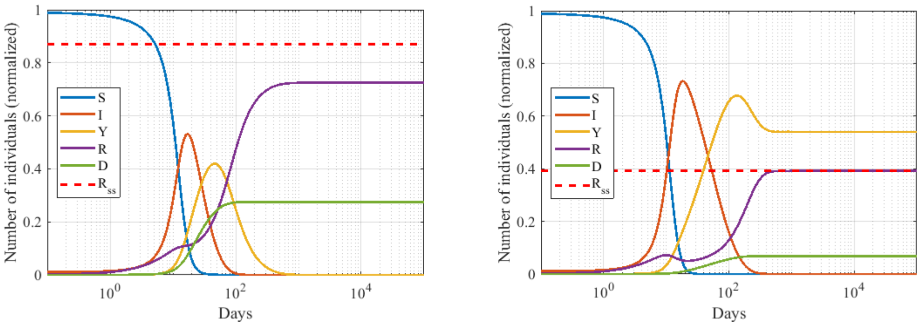


**Fig 2.** Generic dynamics of the SIYRD model in the absence of mortality of reinfected individuals. Comparison between convergence to a non-endemic (left) and endemic (right) steady-states (note the logarithmic $x-$axis). Parameters are $\beta_{SI} = \beta_{RI} = 0.46$, $\beta_{SY} = \beta_{RY} = 0.046$, $\mu_I = 0.018$ (left) and $\mu_I = 0.00149$ (right), $\mu_Y = 0$, $r = 0.04$ (left) and $r = 0.018$ (right), $v = 0.01$, $\theta = 0.7$ and Hill coefficient $z = 8$. The red dashed line represents $R_{ss} = R^*$. On the left, an example of converge to the non-epidemic steady, $X_1^*$, in which it is not possible to derive the analytical expression of the steady state ($R_{ss} \neq R^*$). On the right, convergence to the pure endemic steady state, $X_3^*$, guarantees $R_{ss} = R^*$. Initial conditions: $(0.99, 0.01, 0, 0, 0)$, $N = 1$.

system, the exact values of $Y^*$ and $D^*$ are still unknown. Mathematically, and assuming $S(I = 0) = 0$, these two scenarios can be expressed as follows:

$$\text{If } D|_{I=0} > 1 - \frac{r}{\beta_{RY}} \rightarrow (R+Y)|_{I=0} < \frac{r}{\beta_{RY}} \rightarrow \begin{cases} Y^* = 0 \\ R^* < \frac{r}{\beta_{RY}} \end{cases} \tag{18}$$

$$\text{If } D|_{I=0} < 1 - \frac{r}{\beta_{RY}} \rightarrow (R+Y)|_{I=0} > \frac{r}{\beta_{RY}} \rightarrow \begin{cases} Y^* \neq 0 \\ R^* = \frac{r}{\beta_{RY}} \end{cases} \tag{19}$$

Therefore, a low primary-infected mortality rate combined with a high reinfected to recovered infection rate (or low recovery rate) would raise the likelihood for condition (19) to be satisfied and hence have an endemic steady state.

### Supplementary file 4: **Application of $S^2$IYRD to COVID-19, 50 years threshold**

This section extends the study of the application of $S^2$IYRD model to COVID-19 to the case in which the age threshold between groups is placed at 50 years, significantly below the threshold (at 80 years) used in the main text.

Table 1 summarizes the empirical values obtained for the four different countries in the main text, which differ in their demographic composition and in their contacts (see S1 Appendix and S2 Appendix for raw data). Though $n_2$ increases monotonically with the decrease in the threshold, its values for Israel and SAR still highlight the youth of the population (using the 50 years threshold). IFR values reflect the heterogeneity between countries, with a relevant increase in the ratio between $\text{IFR}_2$ and $\text{IFR}_1$ as compared to the threshold at age 80. Also, the fraction of contacts within and between groups, synthesized by the values of $M_{ij}$, underline differences in social habits. (See S2 Appendix for additional comparisons between the two thresholds explored.)

**Table 1.** Empirical parameters for the countries in Fig. 1. Updated demographic pyramids (to year 2020) and independent, empirical values of COVID-19 IFR for each country per age group have been used for these estimations. Details on data origin are reported in S2 Appendix.

| | $n_2$ | $\text{IFR}_1$ | $\text{IFR}_2$ | $M_{11}$ | $M_{22}$ | $M_{12}$ |
|---|---|---|---|---|---|---|
| Spain | 0.41 | 0.05% | 5.71% | 7.45 | 1.29 | 2.84 |
| SAR | 0.17 | 0.84% | 8.32% | 5.96 | 0.89 | 1.29 |
| Israel | 0.26 | 0.03% | 3.75% | 6.49 | 1.54 | 1.99 |
| USA | 0.55 | 0.14% | 8.98% | 6.19 | 1.82 | 2.52 |

Fig. 1 summarizes the effect of different vaccinating protocols in the reduction of deaths as a function of the vaccination rate $v$ for the four countries above. In all cases, vaccinating first $G_2$ always reduces mortality more than does any of the other two strategies, and its advantages are quantitatively larger than with the 80 years threshold. The relatively smaller advantage of $G_2$ prioritization in the SAR could have been expected in the light of the small value of the ratio between $\text{IFR}_2$ and $\text{IFR}_1$. Israel stands a the opposite end: a higher $\text{IFR}_2/\text{IFR}_1$ ratio entails a larger advantage in terms of death reduction under $G_2$ priority. USA results resemble those of Israel, though the $G_1$-first strategy performs slightly better. This is caused by the comparable population size of groups $G_1$ and $G_2$, which leads to an earlier start of vaccination in the non-prioritized group - especially relevant under $G_1$-first strategy.

Though increasing the vaccination rate monotonically increases the reduction in the number of deaths, regardless the protocol, the improvement is not linear. For example, it leads to limited increases in Israel and USA for $v \approx 0.75$ when the most vulnerable group is vaccinated first (as an example, Israel $G_2$ reduction of deaths barely grows from 67.1% at $v = 0.75$ to 68.8% at $v = 1$). Such saturation reflects the change from $G_2$ to $G_1$ vaccination. Not as outstanding as in USA and Israel, this $G_2$ slowdown is also measurable in SAR, where one can distinguish an initial fast reduction in the number of deaths until $v \simeq 0.35$, and a slower increase in RD afterwards. In Spain, in contrast, the increase is almost linear throughout all vaccination rates.

The improvement in the reduction of deaths for the $G_2$ group is very significant at high vaccination rates, as for the 80 years threshold. In all cases the second-best

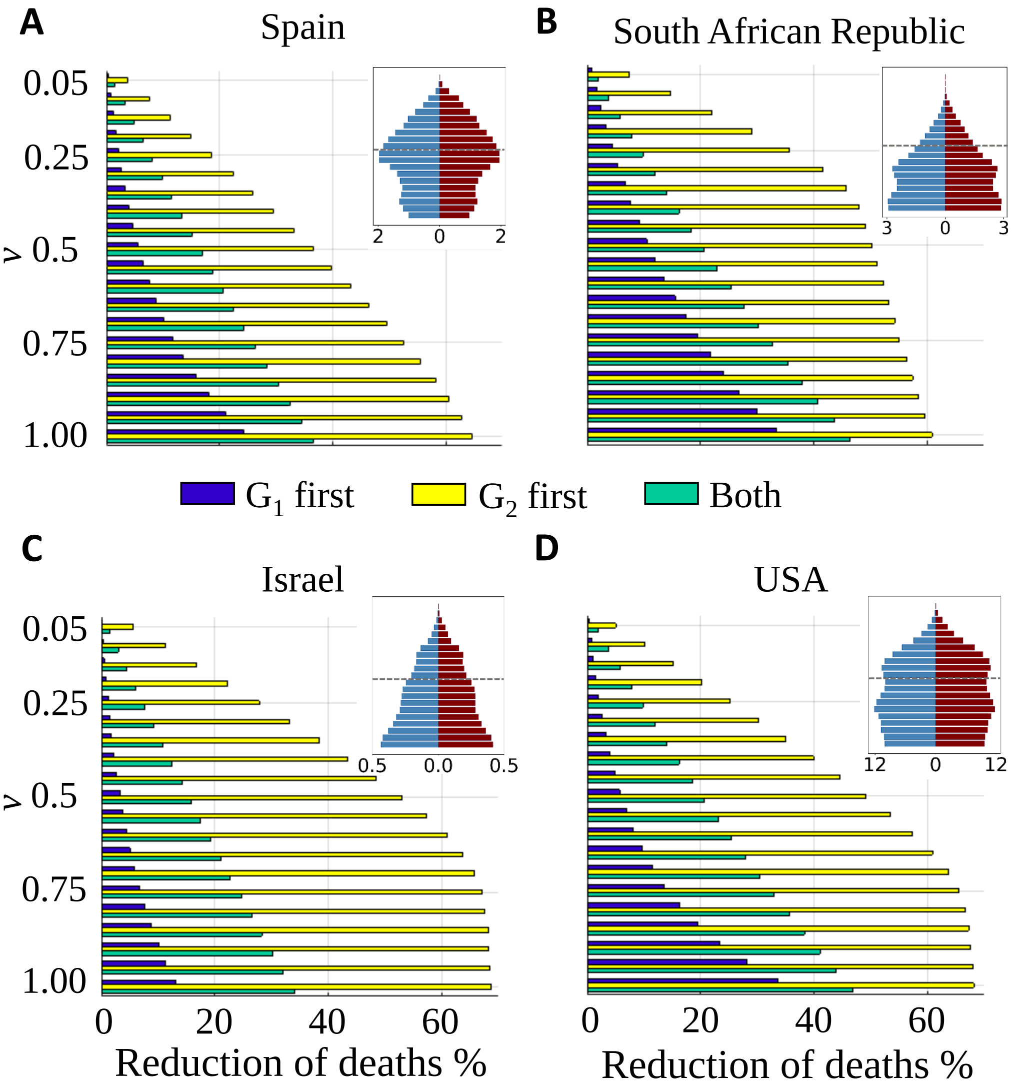


**Fig 1.** Death reduction after one year corresponding to each vaccinating strategy as a function of the vaccination rate. (a) Spain; (b) South African Republic; (c) Israel and (d) USA. Population pyramids are shown as an illustration of the demographic structure; blue corresponds to male population, dark red to female population; numbers in the x-axes of the insets stand for population in millions; vertical bars correspond to 5-year intervals, starting with 0-4 years at the bottom. Demographic data correspond to year 2020 and have been obtained from the Spanish National Institute for Statistics (INE) and from the World population 2019 prospects of UN. IFR measures carried out independently in each of these countries have been used in the simulations; otherwise, model parameters are the same for all cases, with an age threshold at 50 years (indicated as a dashed line in the inset).

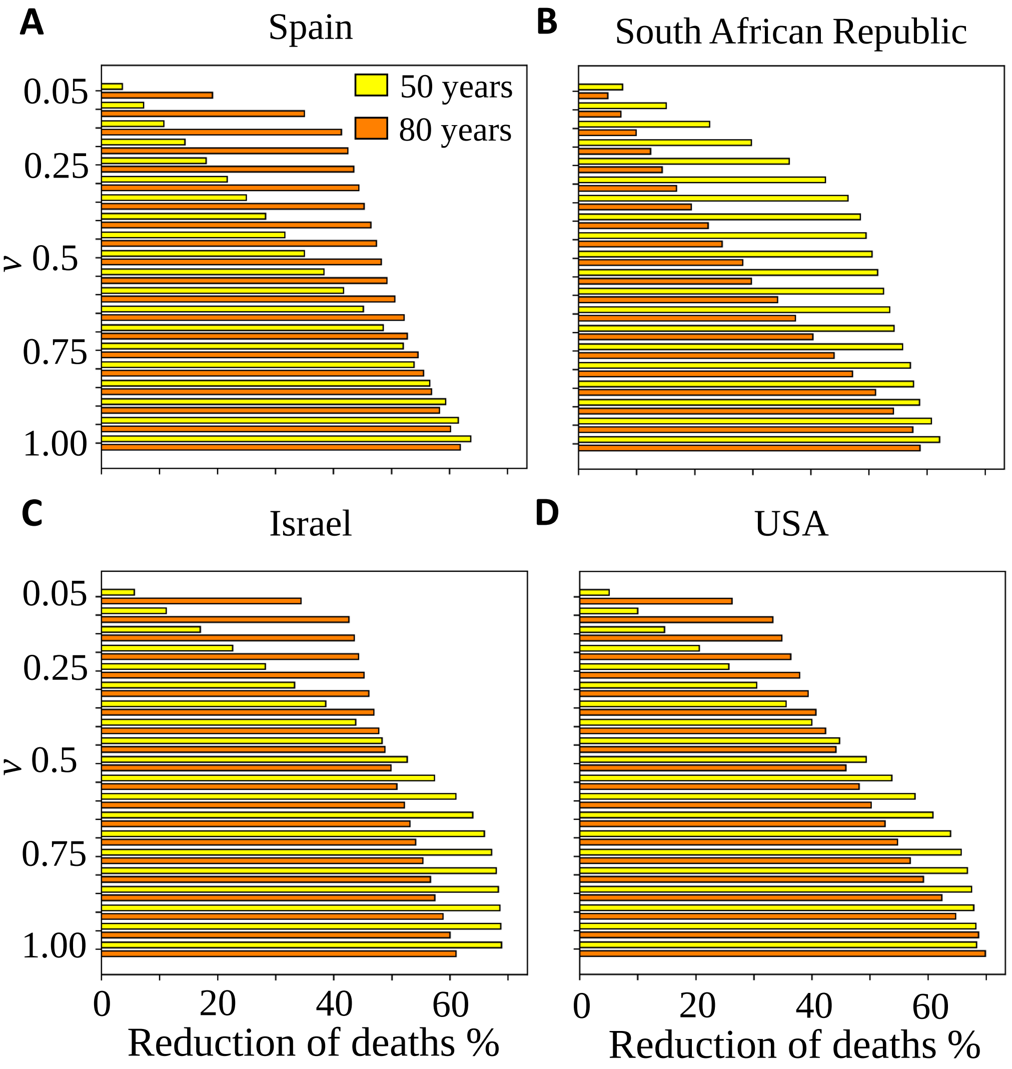


**Fig 2.** Death reduction after one year under application of the $G_2$-first protocol. Each panel compares the reduction of deaths using two different age thresholds as a function of the vaccination rate. (A) Spain; (B) South African Republic; (C) Israel and (D) USA. Yellow bars represent the same data that the yellow bars in Fig. 1.

strategy (simultaneous vaccination) lies far beyond the reduction of deaths achieved by $G_2$ prioritization (by at least 15%). Even in the SAR, which has the smallest difference between strategies at high vaccination rates, the prioritization of the older group avoids at low vaccination rates (e.g. $v = 0.25$) a higher proportion of deaths than $G_1$ prioritization can achieve at high vaccination rates (e.g. $v = 1$). This underscores once more how $G_2$-prioritization allows to attain much earlier in time a death reduction comparable to that eventually attained with any of the other two strategies, due to the early protection of the most susceptible group under the high $\mathrm{IFR}_2/\mathrm{IFR}_1$ ratio of COVID-19.

In case the division into two groups of a population is decided before vaccination roll-out, it is important to clarify that the benefits of one or another divide and protocol vary with the vaccination rate. That is, the optimal split of the population to minimize the number of deaths depends on $v$. In Fig. 2 we compare the effect of dividing the population using thresholds at ages 50 or 80, as the vaccination rate increases. For countries with an old population, like Spain, it is always advantageous to vaccinate first the oldest fraction of the population, at any vaccination rate up to about $v \simeq 1\%$. For very young populations, however, like in the SAR, where the population over 80 is very small, the effects of using such a threshold is effectively equivalent to the simultaneous vaccination of all of the population, without distinction of ages: hence, the benefit for a demographic structure such as that of the SAR is maximized with a threshold at 50 years. For more complex population structures, like in Israel and the USA, the optimal threshold is more sensitive to the vaccination rate: at low rates there is a clear advantage in vaccinating first the elderly, while at high vaccination rates it can be more effective to lower the age threshold in order to cover most of the vulnerable population before vaccinating the younger group. A variety of thresholds for these and other demographic structures can be explored in the model's webpage [1].

#### References

1. Atienza I. $S^2$IYRD Model Simulator; 2021. https://mybinder.org/v2/gh/IkerAtienza/SIYRD/main?urlpath=%2Fvoila%2Frender%2FSimulator.ipynb.
